# Group programmes to improve the skills, confidence and wellbeing of caregivers of children with neurodisability: a systematic review of effects

**DOI:** 10.64898/2026.02.11.26346104

**Authors:** Kirsten Prest, Kirsten Barnicot, Stewart Drew, Catherine Hurt, David Nicklin, Angela Harden, Michelle Heys

## Abstract

**Background:** Caregiver skills training programmes are well-researched in the fields of autism and intellectual disability, but children with motor disorders such as cerebral palsy remain underrepresented despite their high prevalence. These caregivers face unique challenges, and group programmes may provide family-centred care through information provision, problem-solving and peer support.

**Methods:** Systematic searches of five databases (CINAHL, Medline, Embase, PsychINFO and ERIC) were conducted for interventional studies of group programmes aiming to improve the skills, confidence and wellbeing of caregivers of children with neurodisability focusing on motor disorders. Data were extracted on study and intervention characteristics and outcomes. Risk of bias was assessed, effect sizes calculated, and results summarised descriptively using forest plots.

**Results:** Of 6093 studies identified, 21 studies met inclusion criteria (nine randomised-controlled trials, two quasi-experimental and ten pre-post designs). Most reported on programmes developed in resource-constrained settings and addressed caregiver skills, coping strategies, or health-promoting behaviours. Outcomes were grouped according to caregiver wellbeing, caregiver skills and confidence, and social support and family functioning. Child outcomes were reported separately. Most caregiver outcomes showed positive effects, though most studies had high risk of bias due to self-reported outcomes and lack of blinding of intervention allocation and outcome measurement.

**Discussion:** Group-based training programmes show promise for improving caregiver skills and wellbeing. Clinicians and stakeholders in high-income countries may learn from these innovations in low-resource settings. Future research should strengthen protocol reporting, address attrition, control for confounding factors, and establish a core set of caregiver-reported outcomes to better capture programme impact.

**Systematic review registration:** PROSPERO registration CRD42024595002

## Background

‘Neurodisability’, often referred to as ‘neurodevelopmental disabilities’, can be described as an umbrella term for a heterogeneous group of long-term health conditions that result in functional difficulties following a neurological cause such as injury to the brain or neuromuscular system (1). This definition is kept intentionally broad to include children without specific diagnoses, however cerebral palsy (CP) is often used as an exemplar condition (2). Cerebral palsy affects 1.6 per 1000 live births in high-income countries (HICs) and an estimated 3.4 per 1000 in low- and middle-income countries (LMICs) (3). The Global Burden of Disease-WHO Rehabilitation Need Estimator Database has suggested a higher global prevalence of 0.9% for children with moderate to severe motor impairments (4).

Almost all children with CP or other neurodevelopmental disabilities affecting motor function present with at least one comorbid condition, with many having multiple and complex diagnoses that require simultaneous management (5). Examples range from autism (6), Attention Deficit and Hyperactivity Disorder (ADHD) (7), sleep problems, gastrointestinal symptoms, behaviour difficulties, pain (8), to visual impairment (9), and epilepsy (10). Managing the multitude of symptoms and accompanying health systems involves a high burden of care for the children’s caregivers (11). Caregivers in this paper refers to the parent of the child or the person primarily involved in caring for the child. Caregivers frequently present with reduced physical and psychological health, which impacts on their Quality of Life (QoL) (12–15).

Family-centred care refers to the way in which families and healthcare professionals work together to make decisions using principles relating to collaboration, negotiation, information sharing, respecting differences, and understanding the family and community context (16). It is well-established that family-centred care is the gold standard for this population to ensure that healthcare professionals work in partnership with families and children (17–19). This contrasts with the biomedical model of disability, assisting healthcare professionals to look beyond the inherent diagnosis of the child (20) and rather at the holistic picture of the family, community and wider society (21). Family-centred care has been found to be significantly inversely associated with parent stress (22). A key component of family-centred care is the provision of information about the diagnosis and specific information about the child, which families have reported to be lacking (23,24). In England and Wales in the UK, the National Institute for Health and Care Excellent (NICE) clinical guidelines for cerebral palsy in those under 25 years state that caregivers of children with cerebral palsy require timely and up-to-date information about their child’s diagnosis, prognosis, expected development, comorbidities, availability of equipment, financial support, social care, educational placements, and transition to adults’ services (25). An identified gap in service delivery is failure to provide specific information about the child, such as clinicians not explaining results of assessments to parents/caregivers, or therapists not explaining what is happening for the child in a session (26).

The Peninsula Cerebra Research Unit (PenCRU) in the UK have proposed a model for working in partnership with families of children with neurodisability to provide clear, accessible, relevant and up-to-date information (27). However, the most significant component of family-centred care moves beyond the pure provision of information. It involves working in partnership to enable parents and caregivers to feel competent to request information and support, and to confidently advocate for their child’s needs. This forms part of parent activation which refers to their ability to manage, coordinate and advocate for their child’s healthcare needs (28).When parents are supported in developing their skills and confidence in these areas, they are able to cultivate a more collaborative partnership with healthcare professionals, resulting in better outcomes for the child (29).

Supporting caregivers to improve their skills and confidence to care for their child with a neurodisability is an example of a low-cost intervention. This is pertinent for low- and middle-income countries (LMICs), where for example, the prevalence of CP is higher due to factors such as reduced access to skilled health professionals both at birth and in the post-natal period, birth asphyxia, neonatal infections, and untreated jaundice (30). Low-cost interventions are also relevant to high-income countries (HICs), given constraints such as the National Health Service (NHS) workforce crisis in the UK (31).

Much of the literature around caregiver skills training lies within the fields of autism and intellectual disability, rather than motor disorders like CP. Within these fields, positive evidence has been found for the effectiveness of caregiver skills training for both caregivers and children (32). Elements of caregiver skills training have been researched in the field of neurodisability. For example, parent-delivered therapy, as opposed to resource-heavy therapist-delivered sessions, has been found to be effective when trusting relationships are built, when parents have strong support, and when all parties are motivated (33). Parent education, including co-designed home programmes and coaching, allows for self-efficacy to be built, along with a willingness to learn. This not only improves therapy adherence and child outcomes, but can also influence parental wellbeing (33,34). It may be particularly valuable to focus on groups for caregivers of children with neurodisability, since these can provide valuable opportunities for peer support and community building through problem-solving, learning from each other and forming a sense of belonging (35).

Although studies and reviews are moving towards a “noncategorical approach” to health services research, whereby mixed groups are included of children with varied long-term health conditions (36), children with motor disorders continue to be excluded from research when the umbrella term of ‘neurodevelopmental’ disabilities is utilised. For example, a recent systematic review on skills training for caregivers of children with neurodevelopmental disorders excluded families of children with motor disorders, focusing almost exclusively on children with autism or intellectual disability (32). Similarly, a scoping review aiming to understand the implications of peer support for families of children with neurodevelopmental and intellectual disabilities largely found studies relating to autism (37). Having a heterogenous group with different diagnoses can be a barrier to the functioning of peer support groups, particularly when there may be more services and funding opportunities for children with autism (38,39). Although there are examples of successful group programmes focusing on improving the skills, confidence and wellbeing of caregivers of children with diverse needs (40,41), this systematic review focuses on children with physical disabilities that have a neurological cause. This is an attempt to address the underrepresentation of these families in neurodevelopmental research.

Systematic review registers revealed no prior or ongoing reviews with the same objective, although several related reviews were identified (Table 1). Only one review (He et al. (42)) focused specifically on group-based programmes, many of which are highly relevant to this review although studies were limited to LMICs and covered a broad range of diagnoses. The review conducted by Whittingham et al. (43), focused on improving children’s behaviour and parenting style, which differs from the aims of this review to explore programmes that improve caregivers’ knowledge and skills in how to care for their child, and themselves. The reviews conducted by Irwin et al. (44), Branjerdporn et al. (45) and Poojari et al (46) focused on exploring important elements of family-centred care and peer support for this population of interest, but did not focus on group delivery. Novak-Pavlic and colleagues’ ongoing systematic review (47) is also relevant for this study, however, there remains a broad focus on all diagnoses without extracting specific data relevant to children with neurodisability, particularly motor disorders such as cerebral palsy. Finally, Reichow and colleagues’ systematic review (32) aimed to review skills training programmes for caregivers of individuals with neurodevelopmental disorders. However, the review excluded children whose primary diagnosis was a motor disorder, such as cerebral palsy whereas the systematic review described in this paper aims to focus on those children who were excluded.

**Table 1:** Summary of relevant systematic reviews focusing on caregiver programmes aiming to improve outcomes such as wellbeing, skills, and confidence.

| **Reference**<br>**Author (Date) Title** | **Aim** | **Population** | **Intervention examples** | **Date of**<br>**most**<br>**recent**<br>**search** | **Differences from this**<br>**review** |
| --- | --- | --- | --- | --- | --- |
| **Irwin et al. (2019)** A systematic review and meta-analysis of the effectiveness of interventions to improve psychological wellbeing in the parents of children with cerebral palsy. (44) | To evaluate the effectiveness of interventions aimed at improving the psychological wellbeing of caregivers of children with CP. | Caregivers of children with cerebral palsy | Positive parenting intervention, stepping stones triple P, health education intervention, self-management empowerment model intervention, training and support programme, parent-to-parent programme | December 2017 | Did not focus on group delivery.<br>Mainly focused on wellbeing outcomes (only one part of this review). |
| **Whittingham et al. (2011)** Systematic review of the efficacy of parenting interventions for children with cerebral palsy. (43) | To evaluate the efficacy of parenting interventions (i.e. behavioural family intervention and parent training) with parents of children with cerebral palsy (CP) on child behavioural outcomes and parenting style/skill outcomes. | Parents of children with cerebral palsy | Parenting interventions in conjunction with behavioural intervention and oral motor exercises for children with CP and feeding difficulties, the Hanen It Takes Two to Talk programme, and a Functional Communication Training programme for parents. | April 2010 | Did not focus on group delivery.<br>Interventions focused on parenting style and improving children's behaviour. There was an element of caregiver skills outcomes but it did not address caregiver wellbeing. |
| **Branjerdporn et al. (2021)** Efficacy of early interventions with active parent implementation in low-and-Middle income countries for young children with cerebral palsy to improve child development and parent | To determine the efficacy of interventions with active parent implementation for young children with cerebral palsy (CP) to improve child and parent outcomes in low-middle income countries (LMICs). | Parents of children diagnosed with, or at high risk of CP with a mean age less than 60 | General developmental interventions (e.g. community-based rehabilitation, early intervention, education programmes), functional feeding intervention, upper limb training. | December 2020 | Did not focus on group delivery.<br>Interventions were aimed at improving child development and skills, rather than caregiver outcomes.<br>Only focused on LMICs. |
| mental health outcomes: a systematic review. (45) |  | months at study entry |  |  |  |
| **Novak-Pavlic et al. (2022)**<br>Effectiveness of Interventions to Promote Physical, Psychological, and Socioeconomic Well-being Outcomes of Parents of Children With Neurodevelopmental Disabilities: Protocol for a Systematic Review. (47) | To evaluate the effectiveness of interventions aiming to improve the physical, psychological, or socioeconomic well-being of parents of children with neurodevelopmental disabilities when compared to usual care or no care. | Parents of children with neurodevelopmental disabilities | Not published yet | January 2022 | Did not focus on group delivery. Although it focuses on similar outcomes, it has a broader focus on neurodevelopmental disabilities which will include the population of interest in this review, but will also include children with autism and intellectual disabilities. |
| **Reichow et al. (2023)**<br>Caregiver skills training for caregivers of individuals with neurodevelopmental disorders: A systematic review and meta-analysis. (32) | To systematically review the effectiveness of caregiver and parent skills training programs, including caregiver-mediated interventions, for caregivers of individuals with neurodevelopmental disorders | Caregivers of children with neurodevelopmental disabilities (excluding children with motor disorders like CP) | CBT, Stepping Stones Triple P, support groups, Parent Education and Counselling, Hanen Early Language programme, stress management training, positive parenting with video feedback, DIR/Floortime, Joint attention | July 2021 | Did not focus on group delivery. Excluded children with motor disorders like CP. |
| **Poojari et al. (2024)** Effect of family-centered care interventions on well-being of caregivers of children with cerebral palsy: a systematic review. (46) | To synthesize existing evidence on the effectiveness of Family-centred care (FCC) interventions in improving the well-being of caregivers of children with cerebral palsy (CP), and identify the key components of such interventions that are most commonly practiced and deemed effective. | Primary caregivers (>18 years), providing care for children with or at risk of CP at any severity (any level of GMFCS) up to 18 years. | Nutrition programme, stepping stones triple P, parenting acceptance and commitment therapy, parent coaching, conductive education, individualised programmes, livelihood activity programme, community-based rehabilitation, peer-delivered intervention (LEAP-CP). | April 2024 | Did not focus on group delivery.<br>Mainly focused on wellbeing outcomes (only one part of this review). |
| **He et al. (2024)** Group-based caregiver support interventions for children living with disabilities in low- and middle-income countries: Narrative review and analysis of content, outcomes, and implementation factors. (42) | To explore: a) content and composition of current group-based caregiver support and education interventions implemented in LMICs; b) impact of these interventions on caregiver and child outcomes; c) barriers and enablers to implementation of these interventions in LMICs. | Caregivers of children with disabilities in LMICs | Education programmes, parent training, self-help groups, specific programmes to support parents of children with autism, Down Syndrome, Congenital Zika Syndrome, Triple P, psychological support intervention, WHO CST | April 2022 | Only focused on LMICs. Covered a broad range of diagnoses. |

## Objectives

This review aims to answer the question; ‘Do group programmes aimed at caregivers of children with neurodisability improve caregiver wellbeing, skills and confidence?’

The question can be framed using PICO:

- Population: caregivers of children with neurodisability (focusing specifically on physical disabilities)
- Intervention: group programmes
- Comparisons: (if any) no treatment or usual care
- Outcomes: relating to the skills and confidence required to care for their child along with outcomes relating to their own wellbeing.

The review objectives are to:

1. Understand the types of group interventions that focus on caregiver skills training, increasing knowledge and confidence and improving wellbeing for caregivers of children with neurodisability.
2. Systematically review the evidence on the caregiver group programmes described above, to determine pre-post changes and on caregiver outcomes such as confidence/ self-efficacy, empowerment, wellbeing, health literacy and quality of life.

## Methods

The review is registered with the PROSPERO database for systematic reviews and has been reported in accordance with the guidelines outlined in The Preferred Reporting Items for Systematic Reviews and Meta-Analyses (PRISMA) (48).

### Study Selection

#### Types of Studies

Randomised controlled trials (RCTs), quasi-experimental studies and observational studies were included in this review. The rationale for not solely including RCTs is that there have not been many completed in this field, particularly as programmes for parents/carers of children with neurodisability, for example cerebral palsy, have been under researched in comparison to children with autism or other developmental disabilities (32). Including other designs in the study allowed the review to describe more fully the current scope of the evidence base.

Studies were restricted to those published or translated into English, due to limited resources for translation.

#### Types of Participants

##### Inclusion Criteria

Studies that recruited parents or primary caregivers of a child (<18 years) who have a diagnosis of neurodisability were included. As described in the introduction, the term ‘neurodisability’ in this instance refers to children or young people with a long-term health condition and functional impairments due to a neurological cause. In addition, children and young people in this review needed to have physical difficulties with movement and posture. They may have additional comorbidities, but their primary diagnosis should be a motor disorder due to injury in the brain or neuromuscular system, either present at birth or acquired. Cerebral palsy is an example diagnosis, but other metabolic and genetic disorders can mimic CP. Further examples may include Duchenne Muscular Dystrophy, Rett’s Syndrome, brain injury, spina bifida, Spinal Muscular Atrophy (SMA), and occasionally children are diagnosed with Syndrome Without A Name (SWAN).

##### Exclusion Criteria

Parents/ caregivers of children whose primary diagnosis is linked with behaviour (e.g. autism, Attention Deficit and Hyperactivity Disorder (ADHD), dyspraxia, or Tourette’s syndrome), cognition (e.g. intellectual disability, learning disability), or sensory differences (e.g. blindness, deafness) were excluded, although it was expected that many had these as comorbid diagnoses.

#### Types of Settings

Studies were not excluded based on where they were delivered. We included studies based in a range of settings including, but not limited to, primary care clinics, child development centres, schools, children’s centres, hospices or hospitals.

#### Types of Interventions and Comparators

We included any group intervention or programme aimed at parents or caregivers of children and young people with neurodisability that had a purpose of improving their skills, confidence, empowerment, health literacy, quality of life, physical or psychological wellbeing. Where study design allowed, we included studies comparing these interventions to caregivers who received no treatment or an active control (e.g. given an information leaflet), were on a waiting list, or were receiving care as usual. The intervention had to be delivered to caregivers in a group setting. The programmes could include the children and other members of the family, but there needed to be an element which focused particularly on the skills, confidence and/or wellbeing of the caregivers. Studies were excluded if they only ran one session.

#### Types of Outcome Measures

We aimed to evaluate the effectiveness of the programmes for caregiver outcomes, including quality of life, physical and psychological health, skills, confidence, and/or knowledge (primary outcomes). We also extracted data relating to child and family functioning outcomes (secondary outcomes).

##### Search Strategy

We searched five electronic databases: CINAHL (EBSCOhost), Medline (EBSCOhost), Embase (Ovid), APA PsychINFO (EBSCOhost), and ERIC in October 2024. Reference lists of the reviews described in table 1, along with the included studies were hand searched and checked for eligibility. Potential studies were also found through expert contacts and relevant organisations in the field.

The search strategy was developed with City St George’s, University of London’s librarian for the School of Health and Medical Sciences. Subject headings and key words relating to the population, intervention and outcomes were included in the search. Certain keywords from Novak-Pavlic’s systematic review (47) were used too. Table 2 demonstrates an example search strategy for CINAHL.

**Table 2:** CINAHL search strategy.

|  | Search Terms |
| --- | --- |
| (P) Population:<br>Parents/caregivers<br>of children with<br>neurodisability | **Parents/ Caregivers:**<br>1. (MH "Parents") OR (MH "Fathers") OR (MH "Mothers")<br>2. (MH "Caregivers")<br>3. AB parents or mothers or fathers or carers or caregivers<br>4. 1 OR 2 OR 3<br>**Children with neurodisability**<br>5. (MH "Child") OR (MH "Adolescence") OR (MH "Child, Preschool") OR (MH "Infant") OR (MH "Infant, Newborn") OR (MH "Infant, Low Birth Weight")<br>6. AB child* or infan* or adolescen* or teenage* or "young people" or babies or youth<br>7. 5 OR 6<br>8. (MH "Cerebral Palsy") OR (MH "Hypoxia-Ischemia, Brain, Neonatal") OR (MH "Hypoxia-Ischemia, Brain")<br>9. (MH "Brain Injuries")<br>10. (MH "Motor Skills Disorders")<br>11. AB "cerebral palsy" or AB cp or AB "spastic quadriplegi*" or AB "brain injur*" or AB "Hypoxia-Ischemia" or AB hie or AB "complex neurodisabilit*" or AB "motor disorder" or AB "neurological condition" or AB "neurological disorder" or AB "neurodevelopment* dis*" or AB hemiplegi* or AB diplegi* or AB dyskinetic or AB dystoni* or AB "spina bifida" or AB "muscular dystroph*" or AB Duchenne or AB "Rett's Syndrome" or AB "spinal muscular atrophy" or AB sma or AB "syndrome without a name"<br>12. 8 OR 9 OR 10 OR 11<br>13. 7 AND 12 |
| (I) Intervention:<br>Caregiver group<br>programmes | 14. (MH "Program Implementation") OR (MH "Community Programs") OR (MH "Adult Education") OR (MH "Support Groups") OR (MH "Family Centered Care") OR (MH "Peer Counseling")<br>15. AB program* or service or intervention or educat* or training or treatment or therap* or "support group*" or "peer counseling" or "peer counselling" or peer-to-peer or "peer to peer" or "family centered care" or "family centred care" or "skills training"<br>16. 14 OR 15 |
| (O) Outcomes: | 17. (MH "Confidence") OR (MH "Psychological Well-Being") OR (MH "Empowerment") OR (MH "Health Literacy")<br>18. AB skills or AB confiden* or AB "self efficacy" or AB self-efficacy AB wellbeing or AB "well being" or AB well-being or AB "mental health" or AB psychological or AB "quality of life" or AB qol or AB empower* or AB "health literacy" or AB activation or AB "parent activation" or AB motivat* or AB satisfaction or AB "family functioning"<br>19. 17 OR 18<br>20. 4 AND 13 AND 16 AND 19 |

*MH= indexing term (CINAHL heading), AB= terms in the abstract*

### Selection Process

The results from the literature searches were collated and uploaded onto EPPI-Reviewer (49), which integrated with Zotero referencing manager. EPPI-Reviewer is a web-based software programme for managing and analysing data in literature reviews. Duplicates were removed, and titles and abstracts screened against the eligibility criteria. A second reviewer screened titles and abstracts from 10% of the results to ensure consistency (SD). Any discrepancies were discussed between the two reviewers. Efforts were made to contact study authors if relevant information to determine eligibility was missing.

### Data Collection and Extraction

Data extraction was carried out using a data extraction form developed using guidance from the Cochrane Handbook for Systematic Reviews (50). Study authors were contacted if further information was required. The following data items were collected:

1. General information: Authors details, date published, place of publication, and location.
2. Study characteristics: Study aim, design, start and end dates, duration of participation, and ethical approval.
3. Participants: Description of population (the parents/caregivers and their children’s diagnoses including level of severity and co-morbidities), setting, inclusion and exclusion criteria, methods of recruitment, sampling strategy, randomisation, total number at start of study, baseline imbalances, withdrawals and exclusions, age, sex, ethnicity
4. Intervention: Description of intervention using an adapted version of the Template for Intervention Description and Replication (TIDieR) Checklist (51). Items include rationale or goal of intervention, materials required, activities included, the provider, mode of delivery, location, dosage, and costs.
5. Outcomes: The primary outcomes of interest related to the parent/caregiver. These were grouped into *caregiver wellbeing* outcomes (e.g. anxiety, depression, stress, pain, fatigue, quality of life), and *caregiver skills and confidence* outcomes (e.g. skills, knowledge, self-efficacy, empowerment and adaptation to the child). A third category of caregiver outcomes was formed during the review process that related to *social support and family functioning* as they did not fit easily into the first two groups. Outcomes related to the child were extracted too.
6. Information about the above outcomes: Timepoints measured and reported, outcome definition and whether it was validated, the person measuring, scales (upper and lower limits), sample size, type of outcome (dichotomous or continuous), results (numbers or statistics for dichotomous data and mean and standard deviation for continuous data).

Template data collection forms and data extracted from excluded studies can be made available upon request.

### Risk of Bias

The Cochrane Risk of Bias-2 tool (RoB 2) (52) and the Risk Of Bias In Non-randomised Studies - of Interventions - I (ROBINS-I) (53) tool were used to assess the risk of bias for included studies. The ROBINS-I is a domains-based tool using signalling questions, with guidance provided on the interpretation of judging each domain and overall risk of bias (53).

The risk of bias assessments were carried out by two independent reviewers for each of the study outcomes (KP and either SD or DN). Any disagreements were discussed, and a third reviewer brought in if consensus could not be reached. Reporting bias was assessed by referring to the protocol of the study (if available), to ensure that the pre-specified outcomes were sufficiently reported in the results.

### Data Synthesis and Analysis

A meta-analysis was not appropriate for this review due to the diverse nature of the outcome measures and interventions in the included studies (54). Data synthesis instead followed the ‘Synthesis without meta-analysis (SWiM) guideline’ (55). First, results from all studies were grouped according to three broad categories of outcomes relating to either (1) caregiver wellbeing outcomes, (2) caregiver skills and confidence outcomes, and (3) social support and family outcomes.

Means and standard deviations were extracted for all outcomes in the included studies. Data from continuous outcomes were converted to a Standardised Mean Difference (SMD) (Cohen’s d effect size) with a Standard Error (SE) using slightly varying formulas for independent-groups designs and single-group pre-post designs (56). Effect sizes were interpreted as no effect (<0.2), small (0.2), medium (0.5) and large (>0.8) (57). For the few studies that reported dichotomous outcomes, the pre-post change in proportion was converted to Odds Ratios (ORs) and then to SMDs (58). If results were presented as medians and interquartile ranges, then means and standard deviations were estimated from these (59). Confidence intervals (CIs) for each SMD were calculated.

SMDs were adjusted so that positive values demonstrated improvement in the outcome (e.g. a lower score on a depression scale was adjusted to be a positive value). The SMDs and CIs were then inputted to create a forest plot visually presenting the data under the three caregiver outcome categories (1) caregiver wellbeing outcomes, (2) caregiver skills and confidence outcomes, and (3) social support and family outcomes. The SMDs are presented separately (often within each forest plot) for studies with independent-group designs (RCTs and quasi experimental designs) and studies with single group pre-post designs. Data relating to child outcomes were extracted and descriptively presented in a summary table. In the narrative synthesis of these findings, more weight is given to those with larger numbers and a lower risk of bias (54).

The GRADE approach was applied to the most frequently reported outcomes (five or more studies reporting on one outcome) to assess the confidence in the effect estimates (60). The final GRADE score representing certainty of the evidence was given (‘very low’, ‘low’, ‘moderate’ or ‘high’) depending on the quality of the evidence (study design, risk of bias, inconsistency of results, indirectness of evidence, imprecision and other factors such as publication bias).

Descriptions of the intervention components and mechanisms were framed according to the TIDieR Checklist (51).

Post-protocol, several modifications were made including (1) data were synthesised using the SWiM guideline rather than meta-analysis, with effect sizes converted to SMDs and displayed in forest plots, (2) outcomes were grouped under three domains instead of the two originally specified, (3) child outcomes were additionally extracted and presented descriptively, and (4) the GRADE approach was applied to assess the certainty of evidence for the most frequently reported outcomes.

## Results

### Selection of Studies

A total of 9583 studies were identified during the initial search of databases, including CINAHL (1537), MEDLINE (2439), Embase (3441), APA PsychINFO (1386) and ERIC (666). Additional titles were identified through searching reference lists of similar review topics, included studies and through contacts in the field (114). After duplicates (3490) were removed, 6093 papers remained and were screened based on title and abstract. A second reviewer (SD) screened 10% of potential articles (609) with any disagreements being resolved amongst authors, and at times a third reviewer was consulted when a further opinion was required (MH). A total of 64 studies underwent full-text eligibility screening, resulting in 21 studies being included for this review (see figure 1). Studies were excluded if the children’s primary diagnoses did not explicitly relate to cerebral palsy or similar motor disorders as defined in this review (e.g. intellectual disability, developmental delay or Down Syndrome were excluded). It was also decided that studies would be excluded if their population included caregivers of children with acquired brain injuries that resulted primarily in behavioural and social communication difficulties, particularly when the intervention targeted children’s behaviour.

**Figure 1:**
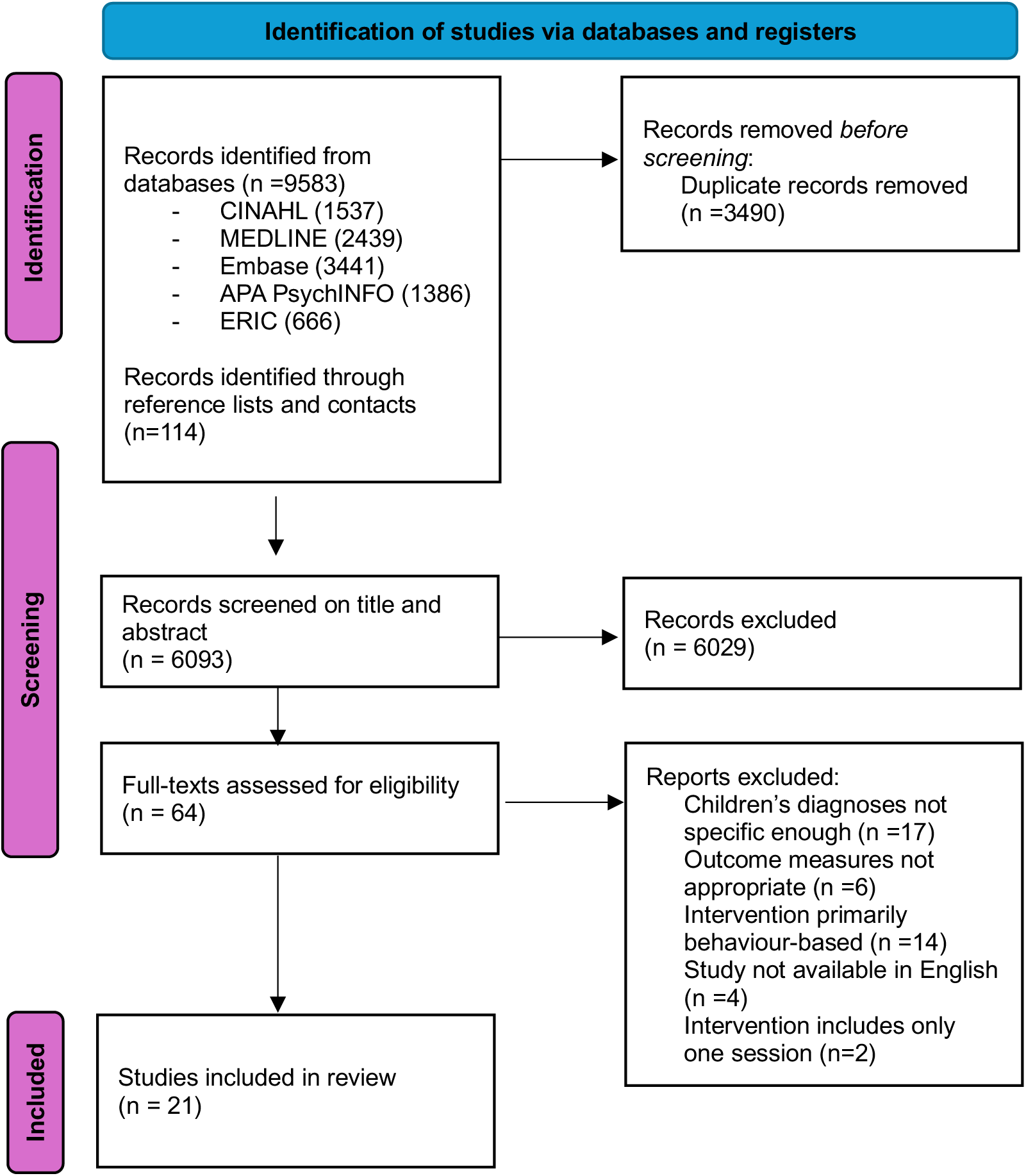
PRISMA ffow diagram.

### Study Characteristics

Out of the 21 studies, including a total of 1491 caregivers, nine were randomised controlled trials, two were quasi-experimental design and ten were single-group pre-post designs (often part of a wider mixed method study) (table 3). Only four studies were set in high-income countries (HICs) (Australia, Hong Kong and United States) with the majority being in low-middle income countries (LMICs). Five of these studies were from South Asia (Bangladesh and India), four from the Middle East and North Africa (Iran and Egypt), one from Europe (Turkey), one from Latin America (Colombia) and six were based in Sub-Saharan Africa (Tanzania, Uganda, South Africa and Ghana). Most studies (n=16) included children with a diagnosis of cerebral palsy and of these, ten studies reported the children being classified as having moderate to severe impairments. The other five studies referenced diagnoses such as neurodevelopmental impairments or special healthcare needs, of which a large proportion of the children had motor difficulties. Thirteen studies reported the mean or median age of the child to be 5 years or under (1 study did not report child age). There were wide ranges in the number of participants included in each study with the smallest studies including 5 or 6 participants and the largest including 251 participants. Twelve studies mentioned that the majority of parents/caregivers were female. Most studies were published between 2018-2025, however two were published considerably earlier (in 1985 and 2000).

**Table 3:** Study Characteristics.

| Study (year) | Country | Study Design | Setting | Participants | Age of parents/caregivers (years) | Age of children | Diagnoses of children and severity | Further information about caregivers presented in study report |
| --- | --- | --- | --- | --- | --- | --- | --- | --- |
| Adams et al. (2011) | Bangladesh | Pre-post intervention study with no control | Child Development and Neurology Unit of the Dhaka Shishu (Children's) Hospital and in three slums of Dhaka | 22 caregivers of children with CP (ages 1- 11years) of moderate to severe disability (GMFCS levels III-V) with feeding difficulties. | Not reported | M (SD) = 3.9 (2.3) years | CP mostly with spasticity and GMFCS level V | 74% of caregivers scored above the threshold for psychological disturbance at baseline |
| Alibakhshi et al. (2024) | Iran | Randomised Controlled Trial | Rehabilitation clinics in Semnan | 52 mothers of children with CP | Intervention group: M (SD) = 32.04 (4.42)<br>Control group: M (SD) = 31.16 (2.98) | Range = 2- 11 years | CP mostly spasticity and GMFCS levels III-V | In order to be included, mothers had to be married and have a primary education level of literacy |
| Al Imam et al. (2022) | Bangladesh | Randomised Controlled Trial | In the community in three rural subdistricts of Sirajganj district | 251 primary caregivers of children with CP under 5 years with the family classified as ultra-poor according to World Bank criteria. | Not reported | M (SD) = 3.5 (1.1) years | CP mostly GMFCS levels III-V | 49% of the children's mothers and 43% of fathers had an education level of secondary and above. |
| Alirahmi et al. (2023) | Iran | Randomised Controlled Trial | Occupational Therapy centres in the city of Ilam | 60 mothers of children with CP who scored highly on a preliminary screening for depression and rumination. | M = 34.13 (no SD) | Not reported | CP | Mothers' education levels ranged from diploma to masters. |
| Asige et al. (2025) | Uganda | Randomised Controlled Trial | 65 villages in Eastern Uganda | 94 caregivers of children/young people with CP | Ranges: 26-45 (46%)<br>46-65 (42%)<br>66-85 (12%) | Range = 2- 23 years | CP with various levels of severity spread across the five levels of the GMFCS and MACS scales. | Most participants were mothers (53%), followed by grandmothers (27%). 51% were subsistence farmers, 68% were married, 28% had secondary education or higher, 71% lived in a rural area. |
| Atilgan et al. (2022) | Turkey | Randomised Controlled Trial | Paediatric rehabilitation Centre in Batman - a major rehabilitation centre in this region | 43 mothers of children with Special Healthcare Needs suffering from chronic low back pain | M (SD) = 34.7 (7.1) | M (SD) = 5 (2.6) years | Mixed diagnoses but mostly CP (37%) followed by Down's Syndrome, Developmental Delay and Spina Bifida | More than half the mothers spent 12 hours or more per day caring for their child. |
| Bourke-Taylor et al. (2018) | Australia | Pre-post intervention study with no control (part of wider mixed methods study) | Mothers recruited from disability organisations and specialised schools supporting students with disabilities | 36 mothers of school-aged children with childhood disabilities | M (SD) = 44.3 (6) | M (SD) = 10.1 (4) years | Mixed diagnoses but 61% had CP and 36% of children needed to be lifted during care | Nearly two-thirds of mothers had professional training, a degree or higher. Most (86%) were partnered. Half (50%) reported that they had been diagnosed with a health condition since their child's diagnosis and half had a mental health diagnosis. |
| Fung et al. (2011) | Hong Kong | Pre-post intervention study with no control | Follow-up clinics at the Department of Orthopaedics and Traumatology, the Duchess of Kent Children's | Six primary caretakers of children with CP | Range = 39-56 | Range = 8-19 years | CP either hemiplegia, dyskinetic CP or quadriplegia and mostly at Special Schools | All female, mostly married and not in paid employment with varying levels of education |
|  |  |  | Hospital, Hong Kong |  |  |  |  |  |
| Hashem et al. (2020) | Egypt | Pre-post intervention study with no control | Outpatient clinic and neurological department at Mansoura University Children's Hospital | 65 caregivers of children with CP | M (SD) = 30.4 (7.5) | M (SD) = 6.19 (4.60) years | CP, with most having a dyskinetic type of CP | 79% were housewives living in rural areas, less than half had secondary education (46%). Most had multiple children and 62% weren't related to their husbands |
| Karim et al. (2021) | Bangladesh | Quasi-experimental design | Early intervention and rehabilitation centre in a rural subdistrict in Northern Bangladesh | 156 caregivers of children with CP identified through the Bangladesh CP Register | Not reported | Intervention group: M (SD) = 4.3 (2.9) years<br>Control group: M (SD) = 11.1 (4.0) years | CP | Most participants lived in mud houses without drinking tap water. Many caregivers were illiterate, and few had secondary education. Most mothers were unemployed; fathers worked in garments, agriculture, or business. |
| McConachie et al. (2000) | Bangladesh | Randomised Controlled Trial | Two settings in the wider study but only the urban groups (near Dhaka) met inclusion criteria | 27 children with cerebral palsy and their mothers | M (SD) = 26.3 (6) | M (SD) = 34.4 (10.9) months | CP, 65% were considered moderate - severe and 54% had severe malnourishment | 19% of mothers had no education and 55% of families had no land. |
| McMillan et al. (2020) | Australia | Quasi-experimental design using a modified stepped wedge cluster | The Royal Children's Hospital (RCH) in Melbourne Australia | 26 parents of children with severe CP aged 12 months to 9 years | Not reported | Range = 12 months - 9 years | CP with GMFCS levels IV or V, 31% had epilepsy and 69% were fully tube fed. | 77% of parents were female and 80% were married. 65% were not born in Australia. 61% were not in paid employment. |
| Mlinda et al. (2018) | Tanzania | Randomised Controlled Trial | CP clinic at Muhimbili National Hospital (MNH) in Dar es Salaam, Tanzania | 110 children with moderate - severe CP under 5 and their caregivers | M (SD) = 30.8 (5.3) | M (SD) = 28 (12.3) months | CP, around half with spasticity and all moderate - severe | 56% of caregivers had an education level of secondary or above. Just over half of caregivers were employed. |
| Muthukaruppan et al. (2020) | India | Pre-post intervention study with no control | A non-governmental organization in South India (ASSA) based in 8 rural locations (blocks) in the District of Tirunelveli, State of Tamil Nadu. | 135 primary caregivers of children with CP who were able to understand Tamil | Not reported | M (SD) = 3 (1.5) years | CP mostly GMFCS levels IV and V | Most caregivers were female |
| Nanyunja et al. (2022) | Uganda | Mixed methods randomised pilot and feasibility trial | Two sites in Uganda - one urban (Kampala) and one rural (Nakaseke) neither of which had existing formal support services for children with developmental disabilities. | 126 caregivers of children with moderate-severe neurodevelopmental impairment | Not reported | Median (IQR) = 9.5 (7.5 - 10.2) months<br>Range = 6 - 12.6 months | Moderate - severe neurodevelopmental impairment | 62% of mothers had secondary education or higher. 70% of fathers had secondary education or higher. |
| Nobakht et al. (2020) | Iran | Randomised Controlled Trial | Occupational therapy clinics in Tehran, Karaj, and Shiraz, (three main cities of Iran) | 91 mothers of children with CP aged 4-12 years | M (SD) = 33.6 (5.8) | M (SD) = 82.62 (29.87) months | CP- GMFCS levels III, IV and V | 35.2% of caregivers had university education and 91.2% were not in paid employment. |
| Smythe et al. (2023) | Colombia | Pre-post intervention study with no control (part of wider mixed methods study) | Community primary school in Cali, Valle del Cauca | 34 caregivers of children with congenital Zika syndrome, CP or toxoplasmosis | Ranges: 15-20 (9%), 21-25 (32%), 26-29 (18%), 30-39 (32%), 40-60 (6%), 61 + (3%) | M (SD) = 26 (11) months | Congenital Zika syndrome (commonly includes spasticity, seizures, eating difficulties, irritability, ocular anomalies, hearing loss, and abnormal neuroimaging). 41% had a diagnosis of epilepsy. | 97% of caregivers were female, 38% of caregivers attended senior school, 15% had job training and none had attended university. 65% of caregivers were unable to work in the previous month. 74% were married or in a civil partnership. 29% owned their home. |
| Tann et al. (2021) | Uganda | Pre-post intervention study with no control (part of wider mixed methods study) | Mulago National Referral Hospital in Kampala | 28 caregivers of infants (under 2 years) with moderate to severe neurodevelopmental impairment | Not reported | M (SD) = 6.7 (0.7) months | Moderate - severe neurodevelopmental impairment. 32% had feeding difficulties, 11% had acute malnutrition and a third were underweight. | Not reported |
| Pilon et al. (1985) | United States | Pre-post intervention study with no control | Cerebral Palsy Clinic at Los Angeles Orthopaedic Hospital | 5 Spanish-speaking parents of children with CP (ages 1-17) | Not reported | Range = 1-17 years | CP | All who attended were women, most of them single parents. |
| van Aswegen et al. (2019) | South Africa | Pre-post intervention study with no control | The Baby Therapy Centre in Mamelodi, a township northeast of Pretoria, | 18 caregivers of children with CP | M (SD) = 32.1 (5.6) | M (SD) = 36.5 (6) months | CP mostly GMFCS level V (66.7%) | All participants were mothers except for one who was a grandmother fulfilling the role of the primary caregiver. 82.4% were unemployed and 61% were single. |
| Zuurmond et al. (2018) | Ghana | Pre-post intervention study with no control (part of wider mixed methods study) | Community settings in 8 districts and 4 regions (Upper East, Greater Accra, Brong Ahafo, Ashanti) in Ghana. | 75 caregivers of children with CP | Ranges: >30 (30%), 30- 40 (45%), >40 (20%) | M (SD) = 3.8 (2.69) years | CP, with half the children considered to have severe CP | 98% of the caregivers were female, with 80% mothers and 16% grandmothers. 67% were married and only half of the biological fathers lived in the same house as their child. 89% described their profession as farming, trading, or a small business such as tailoring. Only 42% had been able to work in the past month. |

CP= Cerebral palsy, GMFCS = Gross Motor Function Classification System, MACS= Manual Ability Classification System

### Intervention Characteristics

The interventions in the 21 included studies tend to be complex interventions with multiple components that were often located in several settings and provided by a variety of professionals, community workers or parents with lived experience (table 4). From an overarching perspective, 15 studies reported on a type of intervention that included learning or practising skills about how to care for their child while providing support to each other in a group. Two of these were focused specifically on feeding and another two reported on the same intervention. Five studies described interventions focusing on providing psychological support and promoting healthy behaviours in a group. One study described a group exercise programme aiming at improving the wellbeing of the parents/caregivers.

**Table 4:**
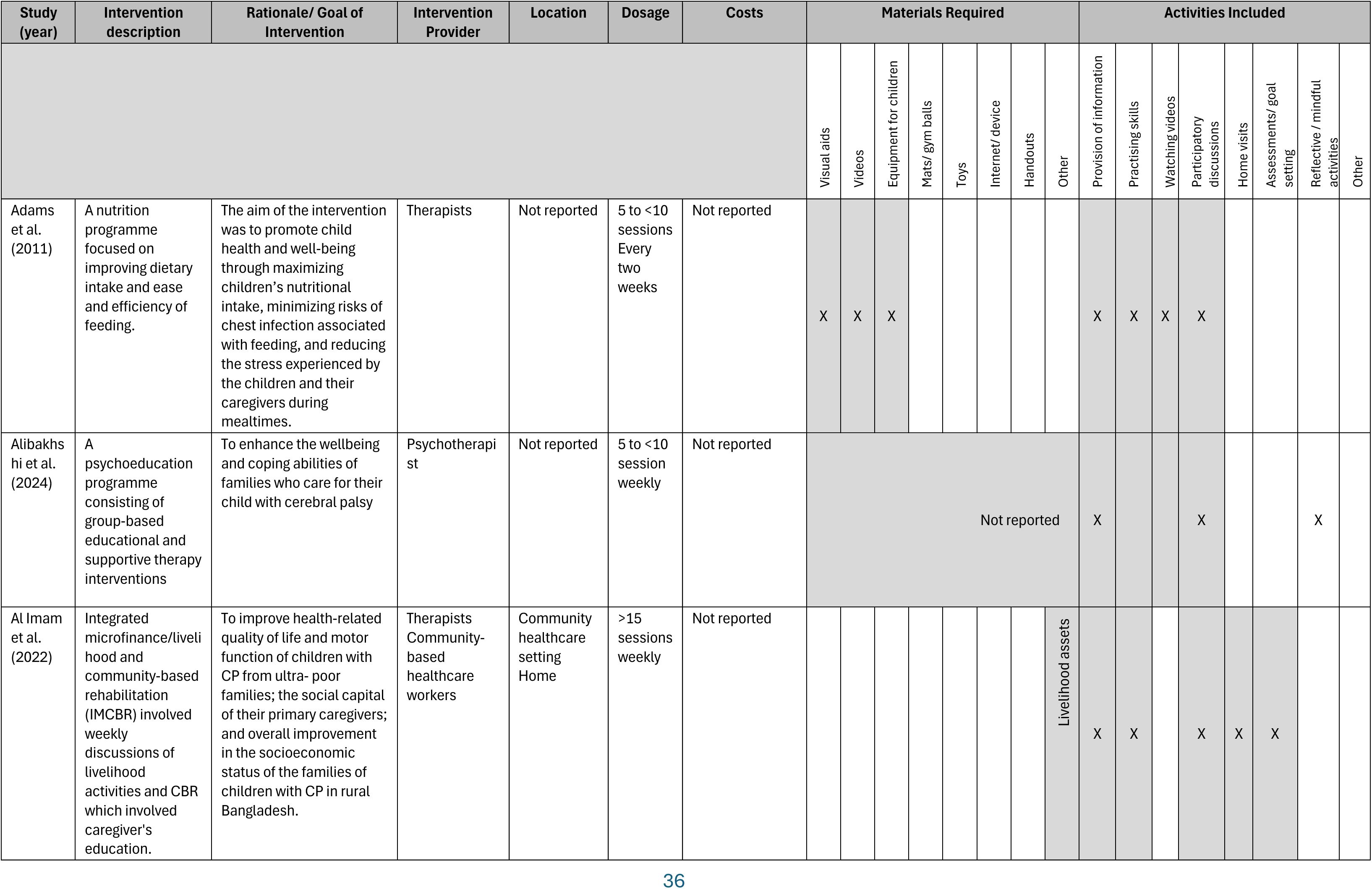
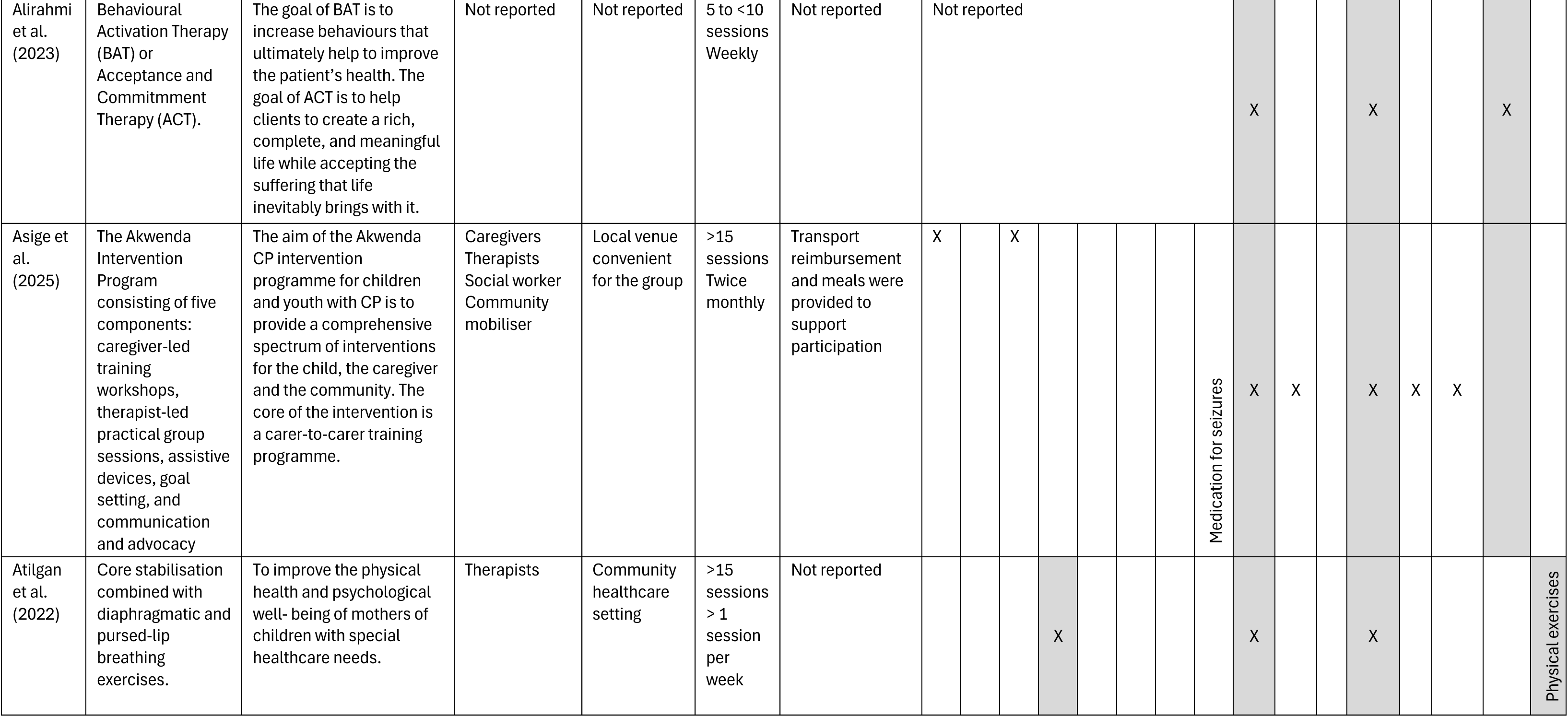
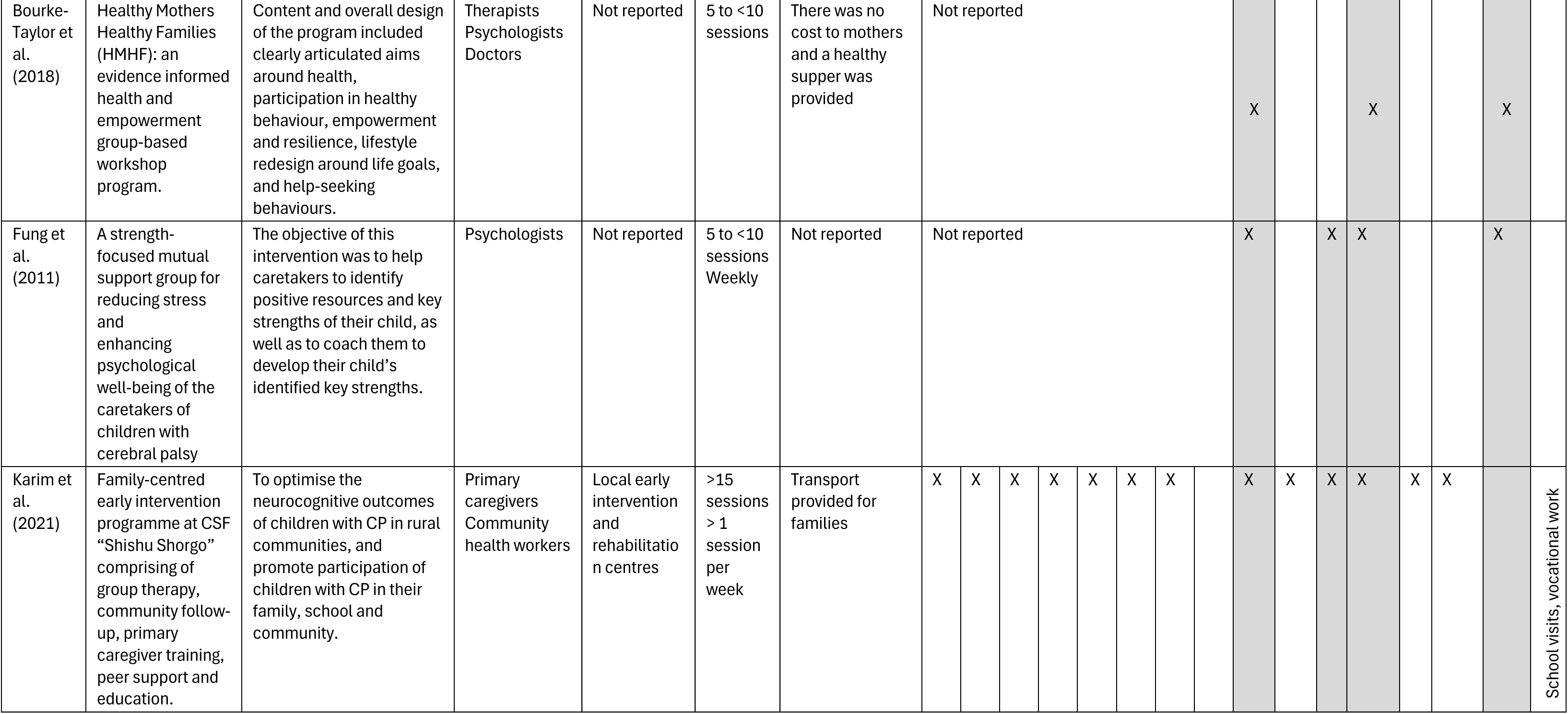
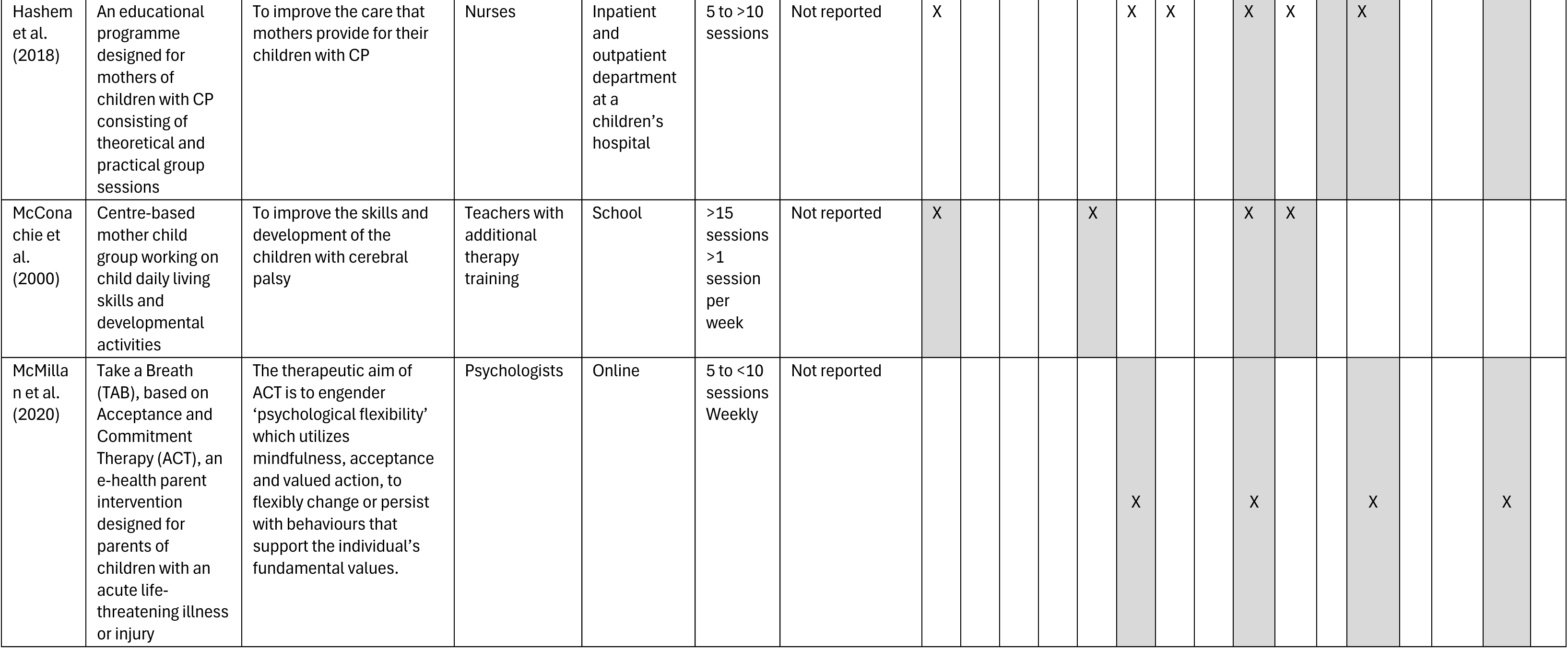
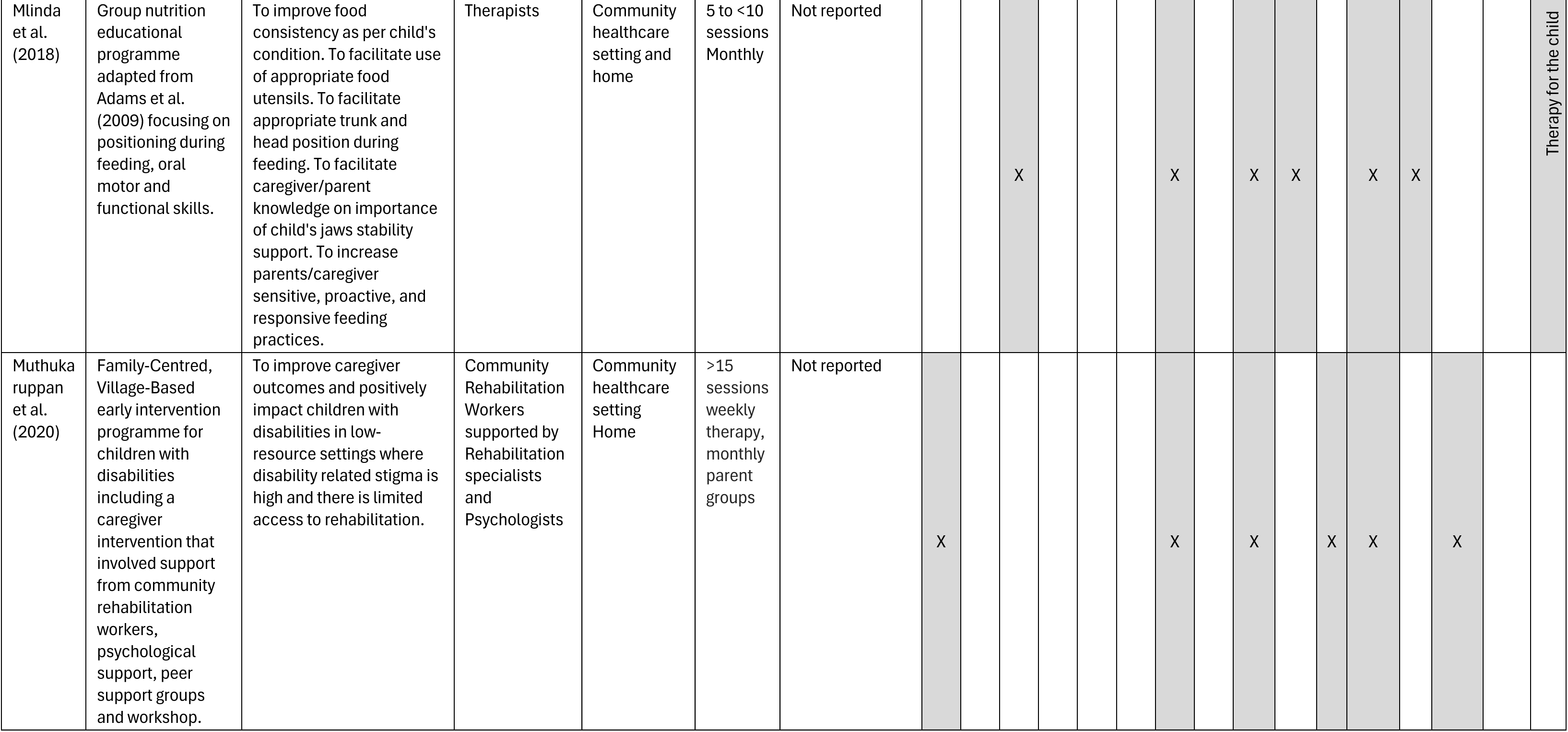
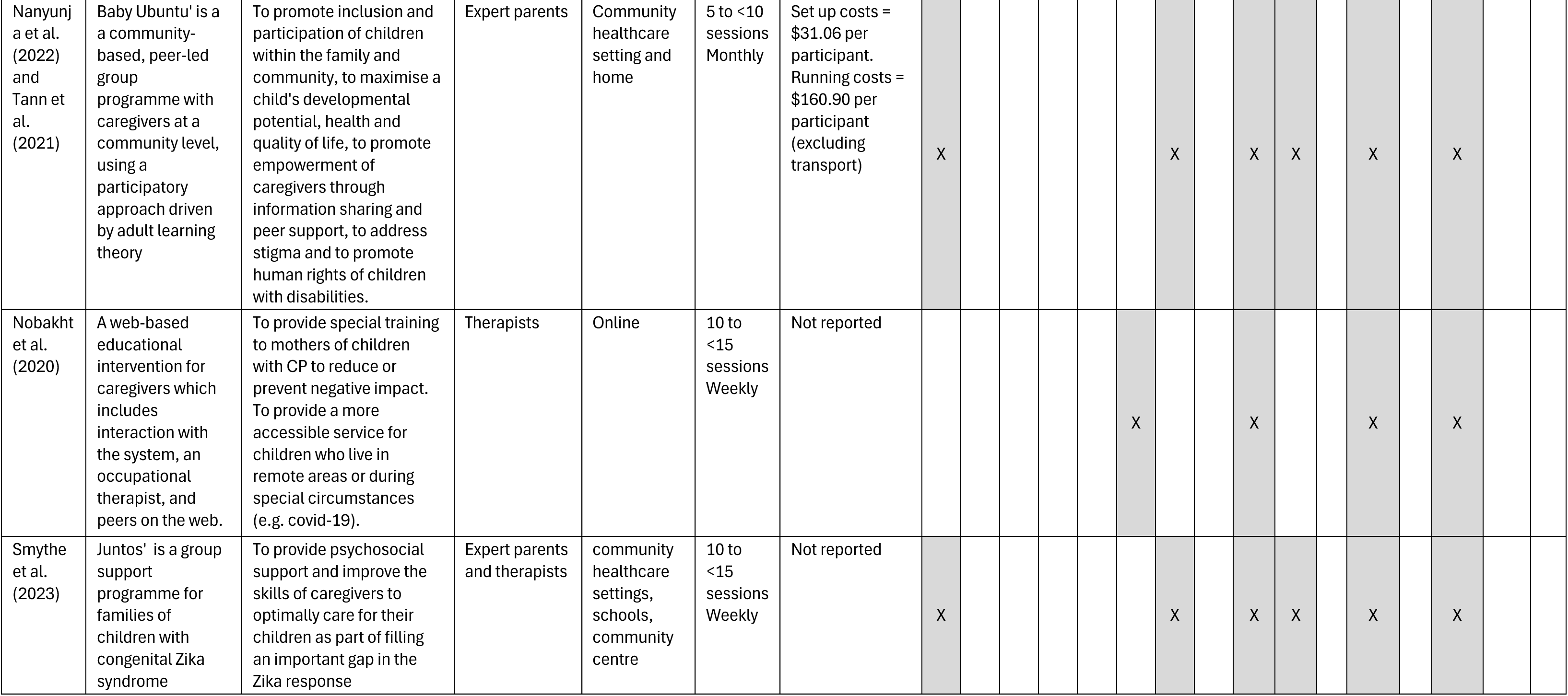
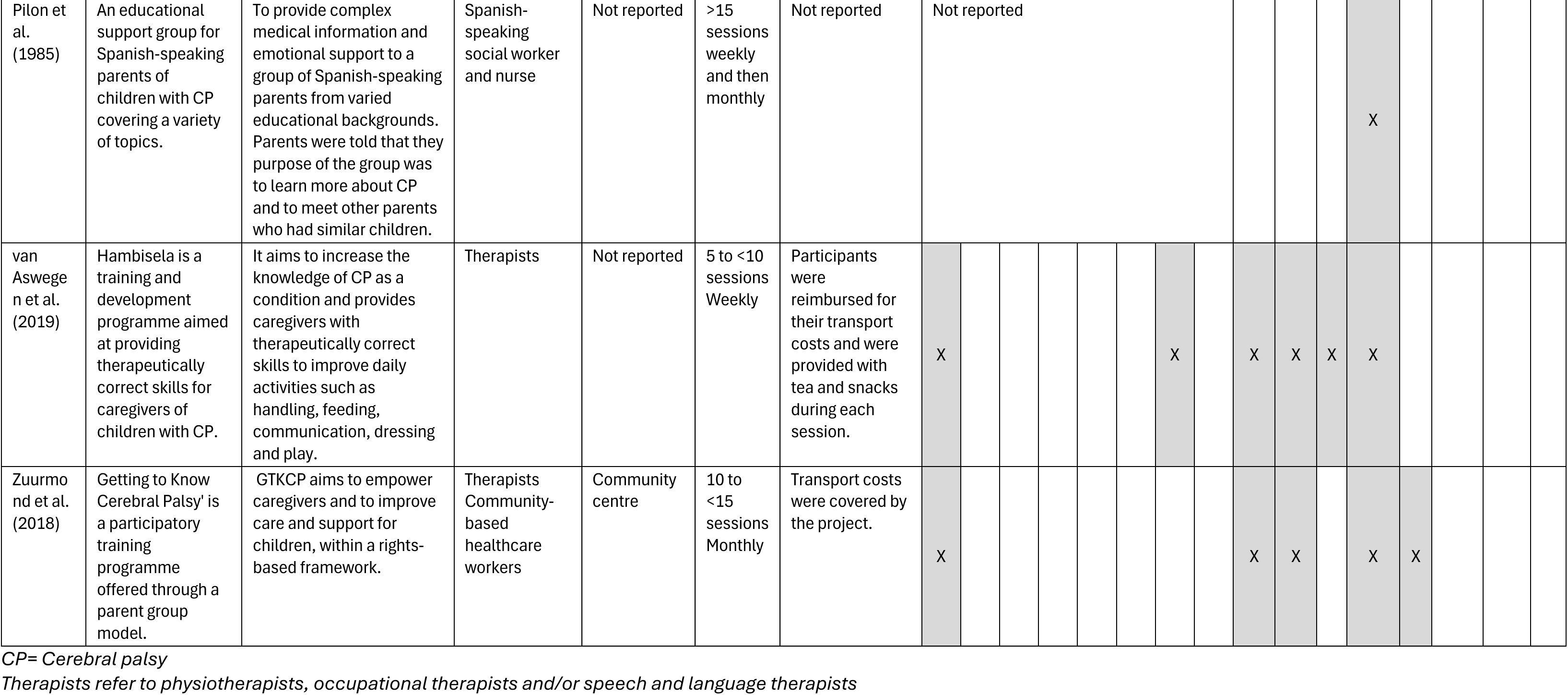
Intervention Characteristics.

Therapists (physiotherapists, occupational therapists or speech and language therapists) were the most common group to deliver the programmes (n=10). Other professionals involved in group delivery were psychologists or psychotherapists (n=5), community-based workers (n=5), nurses (n=2), teachers (n=1), doctors (n=1) and social workers (n=1). Five studies described caregivers themselves delivering the intervention. Most programmes had a combination of individuals described above delivering various aspects of the intervention. Number of sessions included was mostly 5-10 (n=11), followed by 15+ (n=7), with three studies reporting 10-15 sessions. Group sessions were mostly weekly or monthly. Eleven studies reported the intervention location to be in community healthcare settings, local centres or schools. Two programmes were provided online. Few studies reported on the costs of the programme.

The most common activity components of the interventions were provision of information (n=20) and participatory discussions (n=20). Other frequently described activities included; practising caregiving skills (n=12), watching videos (n=5), home visits (n=5), and assessment/goal setting (n=8). Reflective or mindfulness activities were described in all five studies that focused on psychological support or promoting healthy behaviours. The materials required for each intervention were less well reported, however visual aids were frequently mentioned or alluded to (n=11). Equipment for the children, mats, gym balls and toys were mentioned in six studies, but it is likely that these would be utilised in other programmes too. Eight studies described using handouts for caregivers and the four programmes required the use of an internet connection and/or electronic device (either to access the programme online or to watch videos).

The ‘Baby Ubuntu’ community-based peer-led programme described in two of the studies (61,62) currently has a single-blind, effectiveness implementation-hybrid (type II) cluster randomized trial underway in Rwanda (63) and the results will be relevant to this review.

### Risk of Bias

Risk of bias was found to be high for most of the included randomised controlled trials (figure 2). Domain 4 (measurement of the outcome) was found to be at high risk of bias for most of the RCTs due to the nature of the interventions, where blinding was not possible, and the outcome measures themselves which were often self-reported (figure 3). Three RCTs had published protocols which allowed for the comparison between the intended and actual reported results. Most studies had a high proportion of loss to follow up or non-adherence to the intervention.

**Figure 2:**
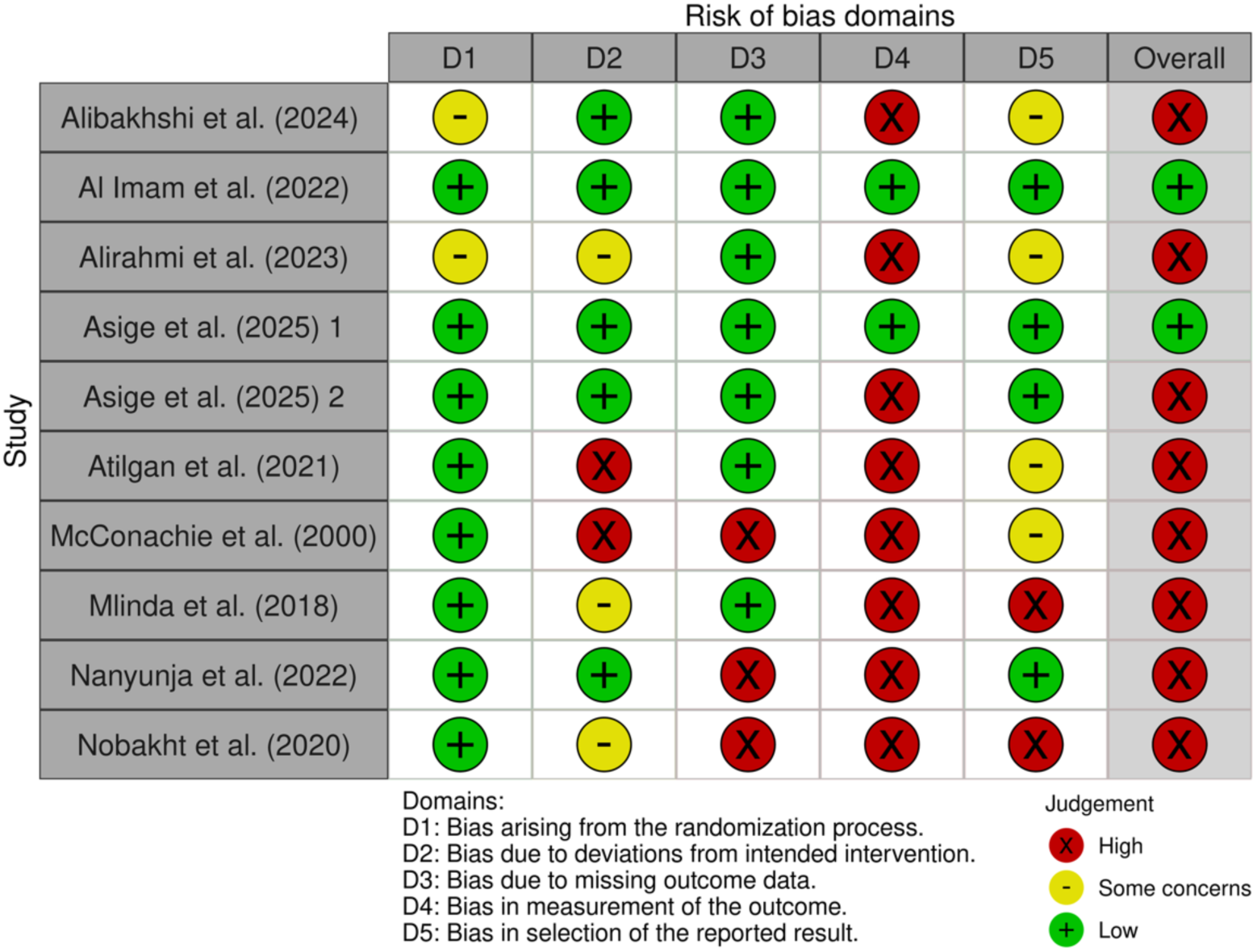
Risk of Bias for included Randomised Controlled Trials. Asige et al. (2025) 1 refers to the outcomes in the study assessed by blinded assessors (e.g. caregiver skills videoed and scored by blinded assessors) while Asige et al. (2025) 2 refers to the self-reported outcomes (e.g. caregiver burden and stress and family functioning).

**Figure 3:**
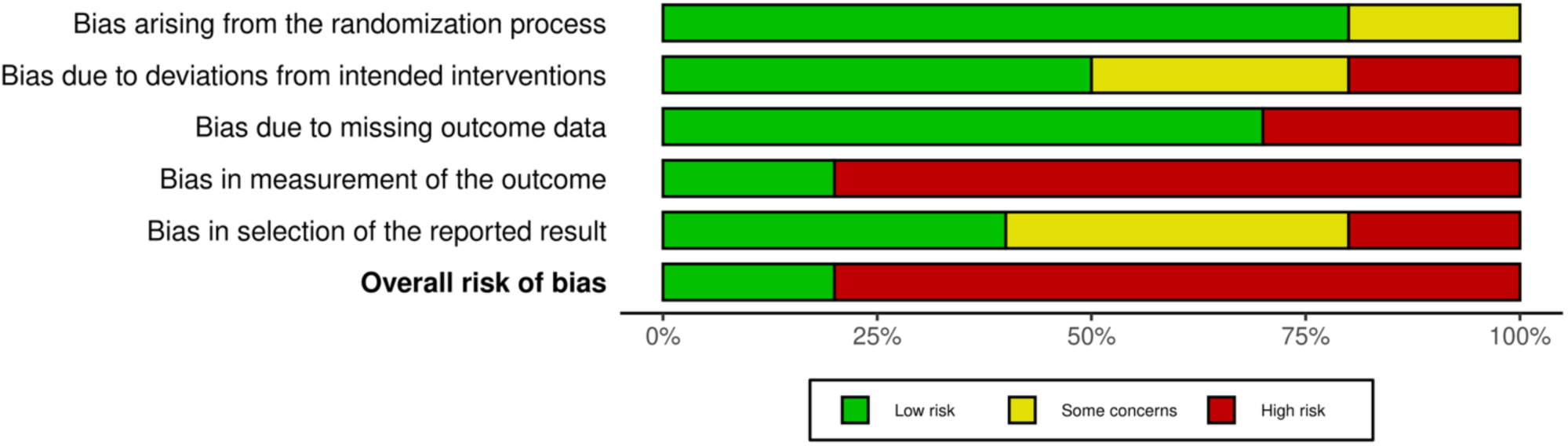
Summary of domains for included Randomised Controlled Trials.

Full ROBINS-I assessments were conducted with three out of a potential twelve non-randomised studies (figure 4). The rest were defaulted to being at ‘critical risk of bias’ due to not being able to evidence any attempt at controlling for confounding. The three that were assessed fully were either at ‘moderate’, ‘serious’ or ‘critical’ risk of bias due to issues with confounding, missing data, or in the measurement of the outcomes.

**Figure 4:**
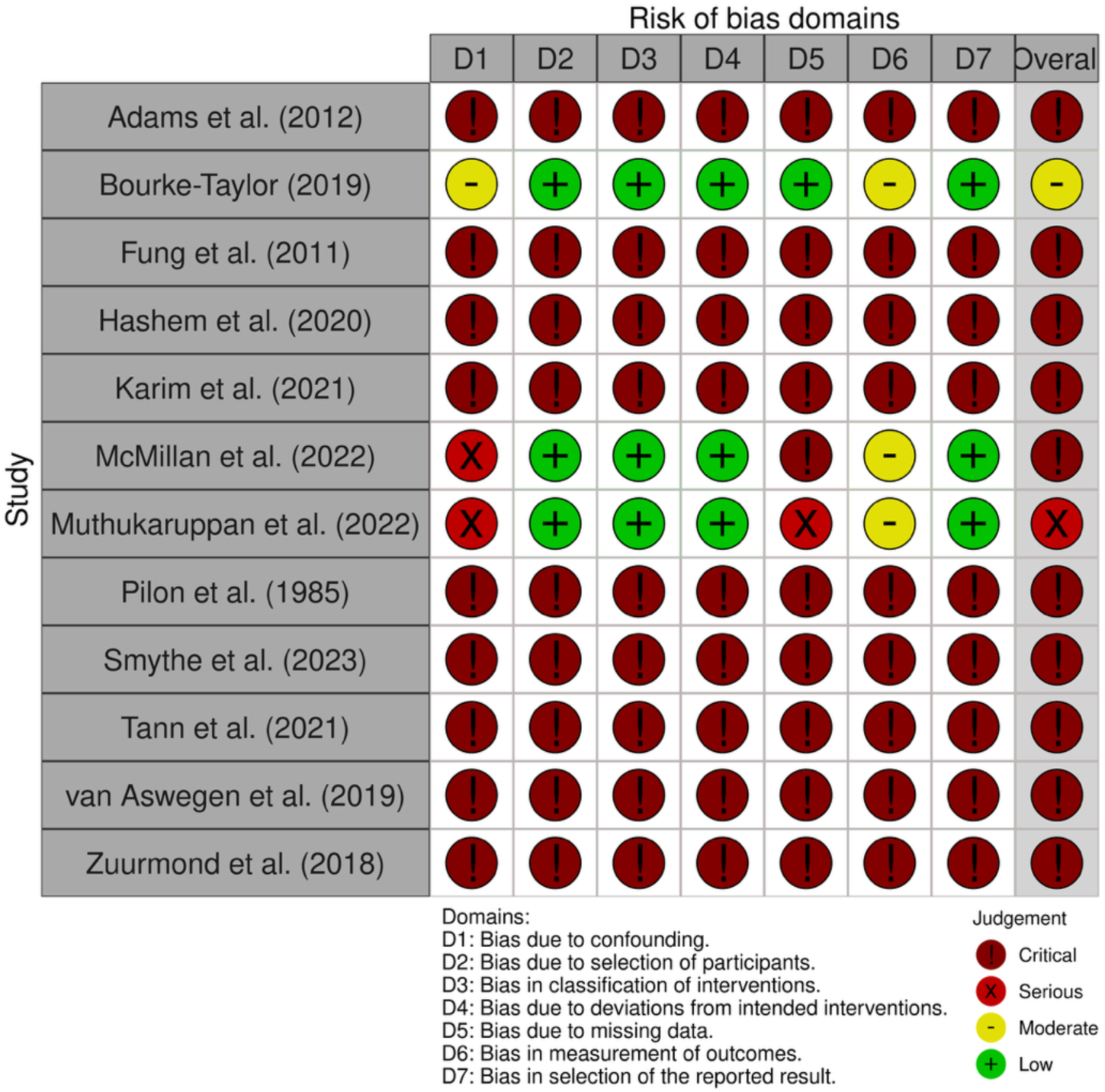
Risk of Bias for included non-randomised studies.

### Caregiver wellbeing outcomes

Of the outcomes relating to caregiver wellbeing (figure 5 and 6), most commonly reported were stress and anxiety (n=18), followed by general wellbeing (including physical activity and mindfulness) (n=15), pain (n=6), depression (n=7), quality of life (n=7), fatigue (n=1) and sleep (n=1). The most statistically significant results relating to improved caregiver wellbeing came from multi-faceted programmes that aimed to improve caregiver knowledge and skills to care for their children and themselves. In contrast, the programmes specifically aimed at improving psychological wellbeing of caregivers were generally smaller studies with higher risks of bias and methodological concerns, with results being more mixed.

**Figure 5:**
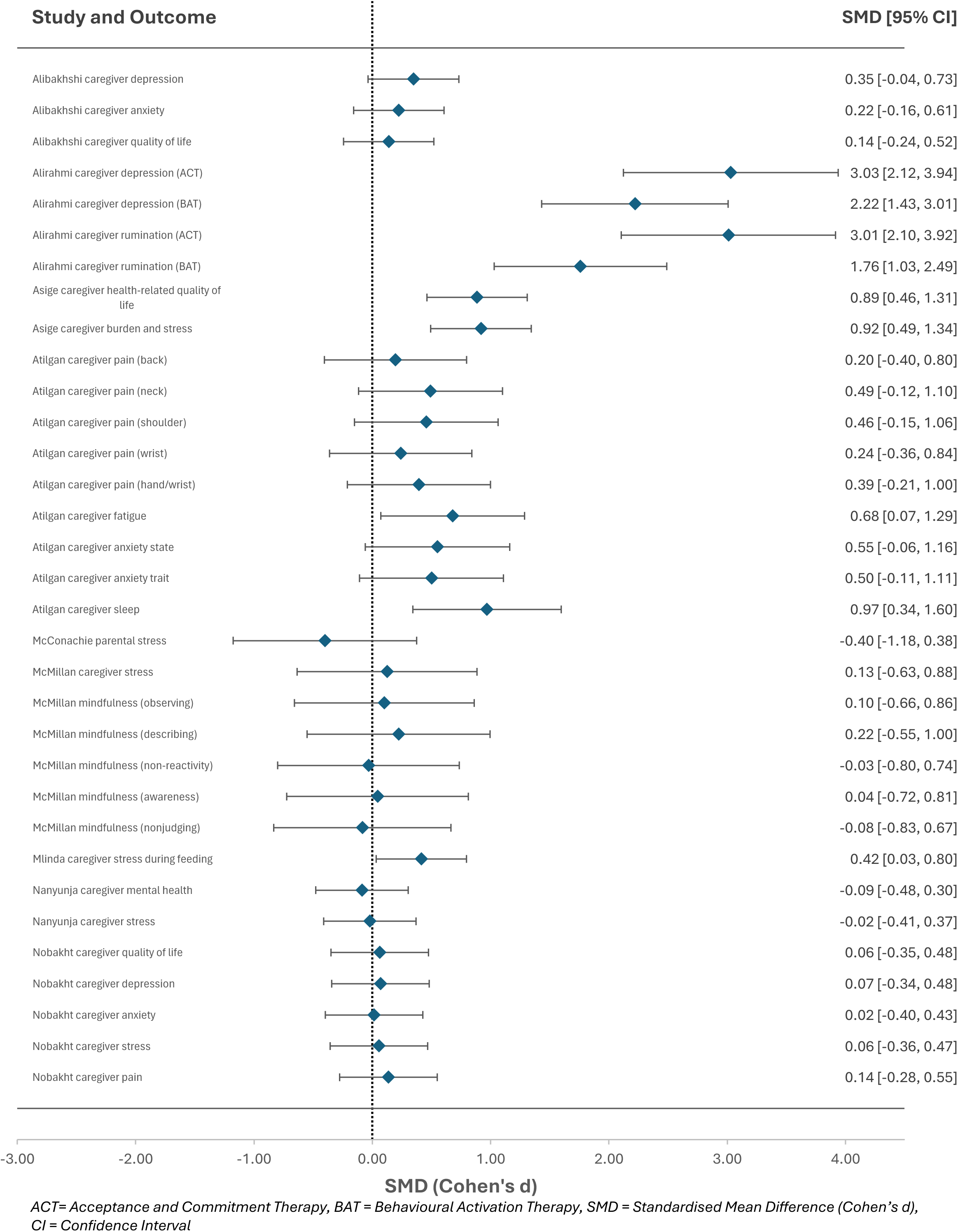
Caregiver wellbeing outcomes for independent group designs (intervention and control groups)

**Figure 6:**
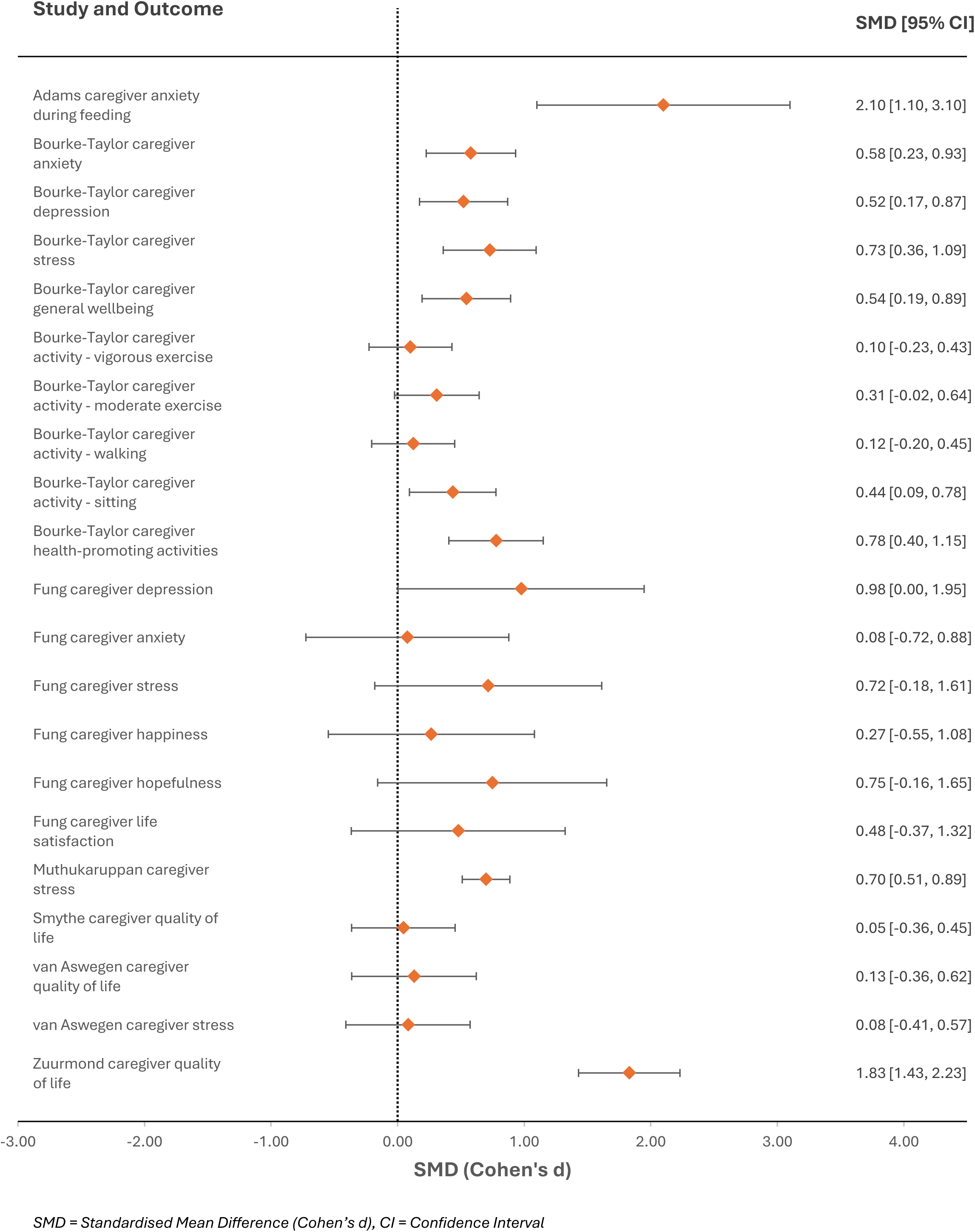
Caregiver wellbeing outcomes for single group pre-post designs (no control)

#### Independent group designs

Although the results from the independent-group studies generally favoured the intervention groups, most are not statistically significant (figure 5). Asige et al. (2025), one of the larger RCTs (n=94) of a multi-faceted programme with a lower risk of bias, found medium to large effects in improving caregiver health-related QoL (d=0.89), and reducing caregiver burden and stress (d=0.92) compared with usual care (64). Another RCT with over 100 participants found a small effect in reducing caregiver stress during feeding following a nutrition programme compared to usual care (d=0.42) (65). Although this is supported by a large effect size in the reduction of caregiver stress in a single-group pre-post study design for the same nutrition programme (66), results should be interpreted with caution as outcomes were unclear, non-standardised and dichotomous. Alibakhshi et al. (2024), an RCT of a psychoeducation programme with over 50 participants found small effects for improving caregiver depression (d=0.35) and anxiety (d=0.22) with no effect found on caregiver QoL (d=0.14) compared to no intervention (67). Another large feasibility RCT of a peer-led caregiver programme with over 100 participants found no effect on caregiver mental health (d=-0.09) and stress (d=-0.02) compared to standard care (61). Karim et al. (2021), a large quasi-experimental study (n=156) on a community-based early intervention programme had positive effects on caregiver depression, anxiety and stress in the short-term, however these were not sustained at follow up. Results were presented descriptively using ordinal data with no means or SDs available, thus this was the only study which could not be included in the forest plot. The improved outcomes with large effect sizes relating to caregiver depression and rumination after participating in Behavioural Activation Therapy or Acceptance and Commitment Therapy compared with usual care (68) should be interpreted with caution, as this was the only study with concerns relating to the randomisation process, and minimal information was available relating to baseline characteristics, loss to follow up or intervention adherence.

#### Single-group pre-post designs

Similar patterns of positive impact are seen in the single group pre-post designs (figure 6), particularly in relation to the reduction of anxiety, depression and stress (69–71). Muthukaruppan et al. (2022), the largest pre-post study (n=135) of a family-centred, village-based early intervention programme with a lower risk of bias than many of the other studies found a medium effect in reducing caregiver stress (d=0.70) (71). A smaller study (n=36) of a health and empowerment group programme with the lowest risk of bias compared to the other pre-post designs, found medium effects for reducing caregiver anxiety (d=0.58) and depression (d=0.52), and improving general wellbeing (d=0.54) and health promoting activities (d=0.78) (69). No negative effects were reported in the single-group pre-post design outcomes.

### Caregiver skills and confidence outcomes

The outcomes in this category were broad (figure 7) and explored caregiver adaptation to the child (including adjusting to the child’s diagnosis and degree of overprotection) (n=6), caregiver skills (often in relation to feeding (n=8), caregiver confidence, self-esteem and empowerment (n=3) and caregiver knowledge about their child’s condition and care (n=5). All outcomes in this category were positive with varying degrees of significance and the largest effect sizes seen in the single-group pre-post study designs. Of the independent group designs, the greatest effects were seen in one of the larger RCTs (n=94), with the lowest risk of bias, in caregiver knowledge (d=1.11), and skills related to dressing (d=0.67) and feeding (d=0.87)(64). Many of the outcomes relating to confidence or knowledge were assessed using non-standardised measures, however two studies used the Family Empowerment Scale (both pre-post single group design but with lower risk of bias compared to others in this review) and both found significant medium effect sizes (69,71).

**Figure 7:**
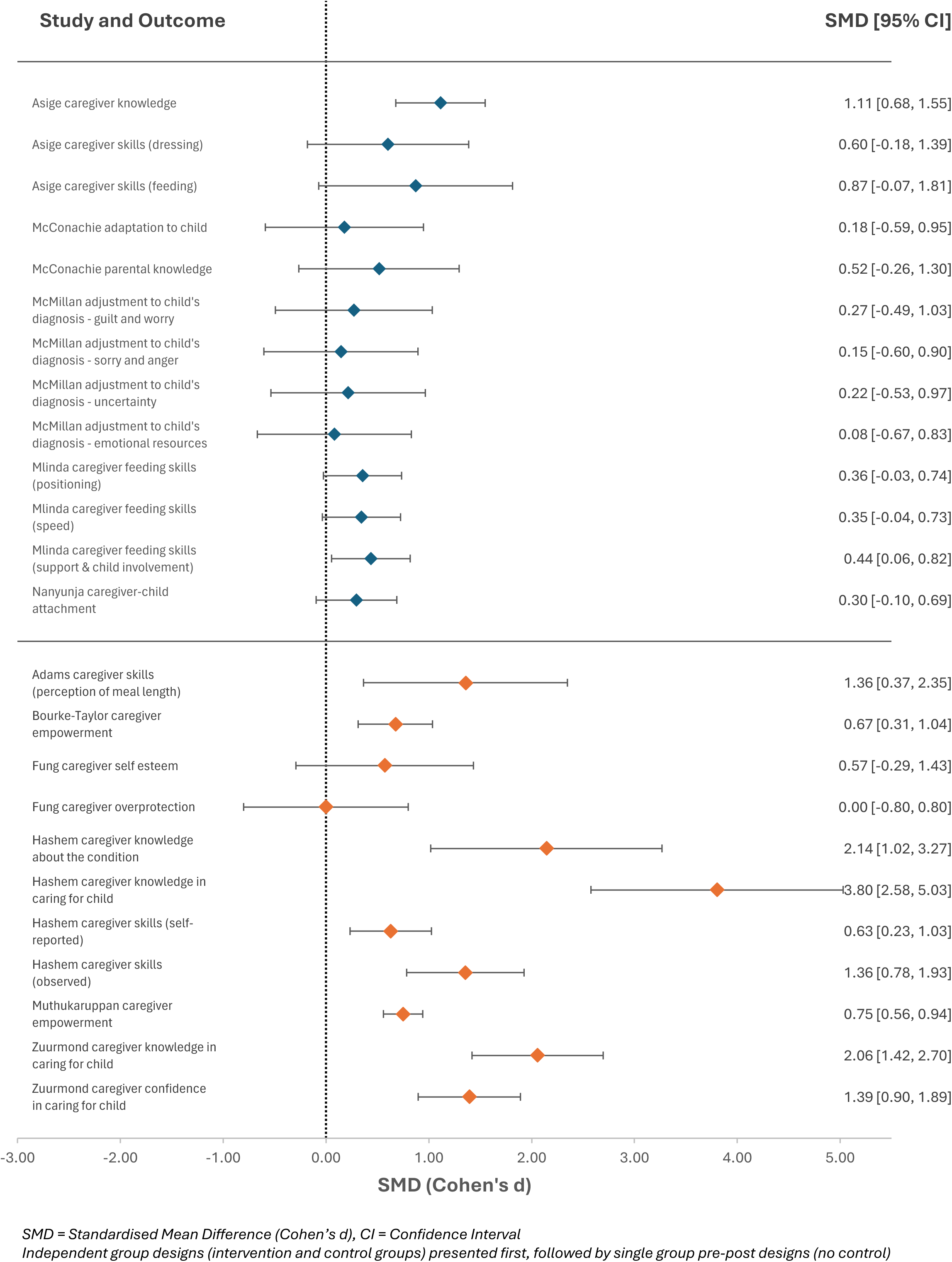
Caregiver skills and confidence outcomes.

### Social support and family outcomes

Outcomes relating to social support, social capital, family quality of life and family functioning were grouped in this category (figure 8). The greatest effects found in this category come from Al Imam et al. (2022), the largest study in this review (n=251), and a cluster-randomised controlled trial with a lower risk of bias compared to most of the other RCTs in this review (72). Compared with care as usual, an integrated microfinance/livelihood and community-based rehabilitation (CBR) programme greatly improved caregiver social capital (d=1.95) and a group receiving CBR without the livelihood component demonstrated greatly improved social capital compared to usual care too (d=1.83).

**Figure 8:**
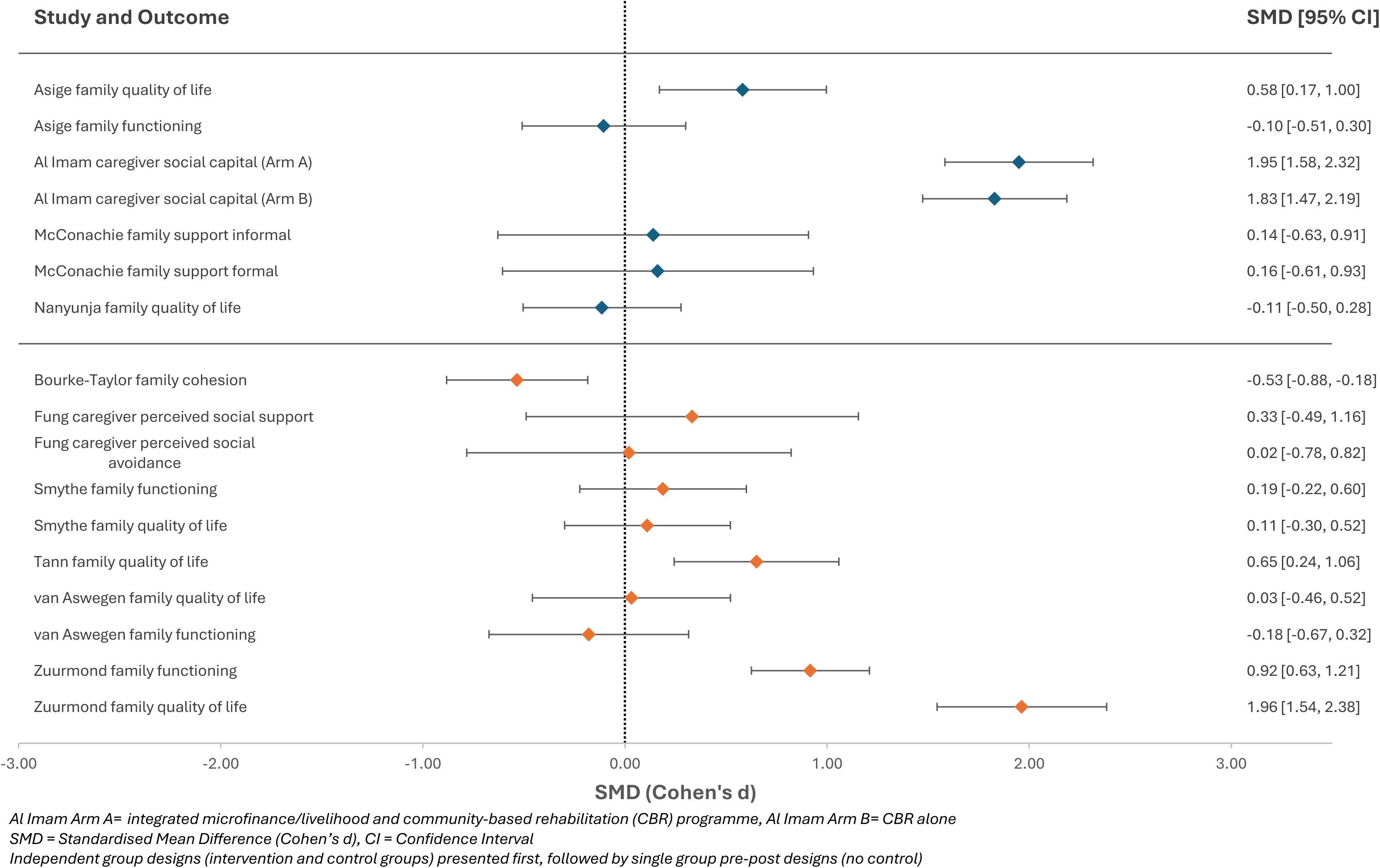
Social support and family outcomes.

Although social support outcomes showed a positive impact, none were significant across the two studies that measured these with both studies either being smaller or at a higher risk of bias (70,73). The most commonly used instrument to measure family quality of life along with family functioning was the PedsQL Family Impact Measure, which was used in six studies, all of which had similar caregiver training intervention components. Mixed results were seen in relation to the impact of these programmes on family quality of life with significant large effect sizes found in three studies (one of which is a large RCT with a lower risk of bias) (62,64,74) and no effect seen in the other three (61,75,76). Family cohesion and family functioning were two outcomes that were negatively impacted in three studies (64,69,76).

### Child-related outcomes

Of the outcomes relating to the child, four studies used anthropometric measures, three studies reported on child quality of life, two studies reported on the child’s feeding skills, and four used measures to assess the child’s motor and cognitive functioning (table 5). The two larger RCTs which were multi-component programmes with the lowest risk of bias reported either small or no effects on child motor function and quality of life, even though large effects were seen in relation to caregiver wellbeing, family quality of life, and social capital as a result of these programmes (72,77). Another large RCT (n=110) reported small to medium effects in improved child feeding skills and mood during feeding as a result of a group nutrition programme compared with usual care (65), with results echoed in a smaller pre-post single group study design on the same programme (66). A larger feasibility RCT (n=126) of a peer-led caregiver programme with a focus on improving caregiver skills found either negative or positive small effects on child growth, motor and cognitive function (61). A smaller pre-post single group study (n=36) on a health and empowerment group programme with a lower risk of bias found small to large effects on child quality of life physical functioning (d=0.48) and psychosocial functioning (d=0.83) (69).

**Table 5:**
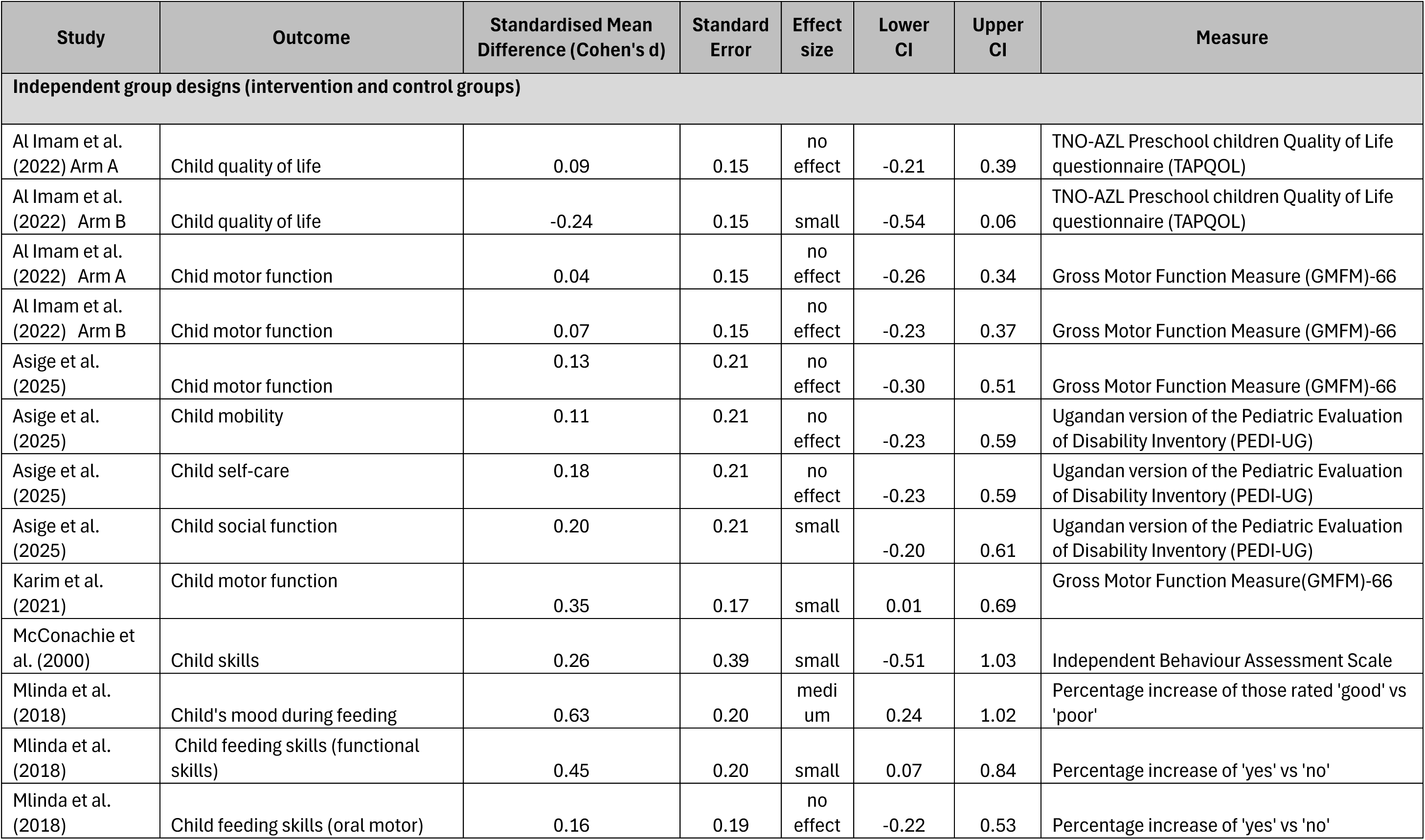
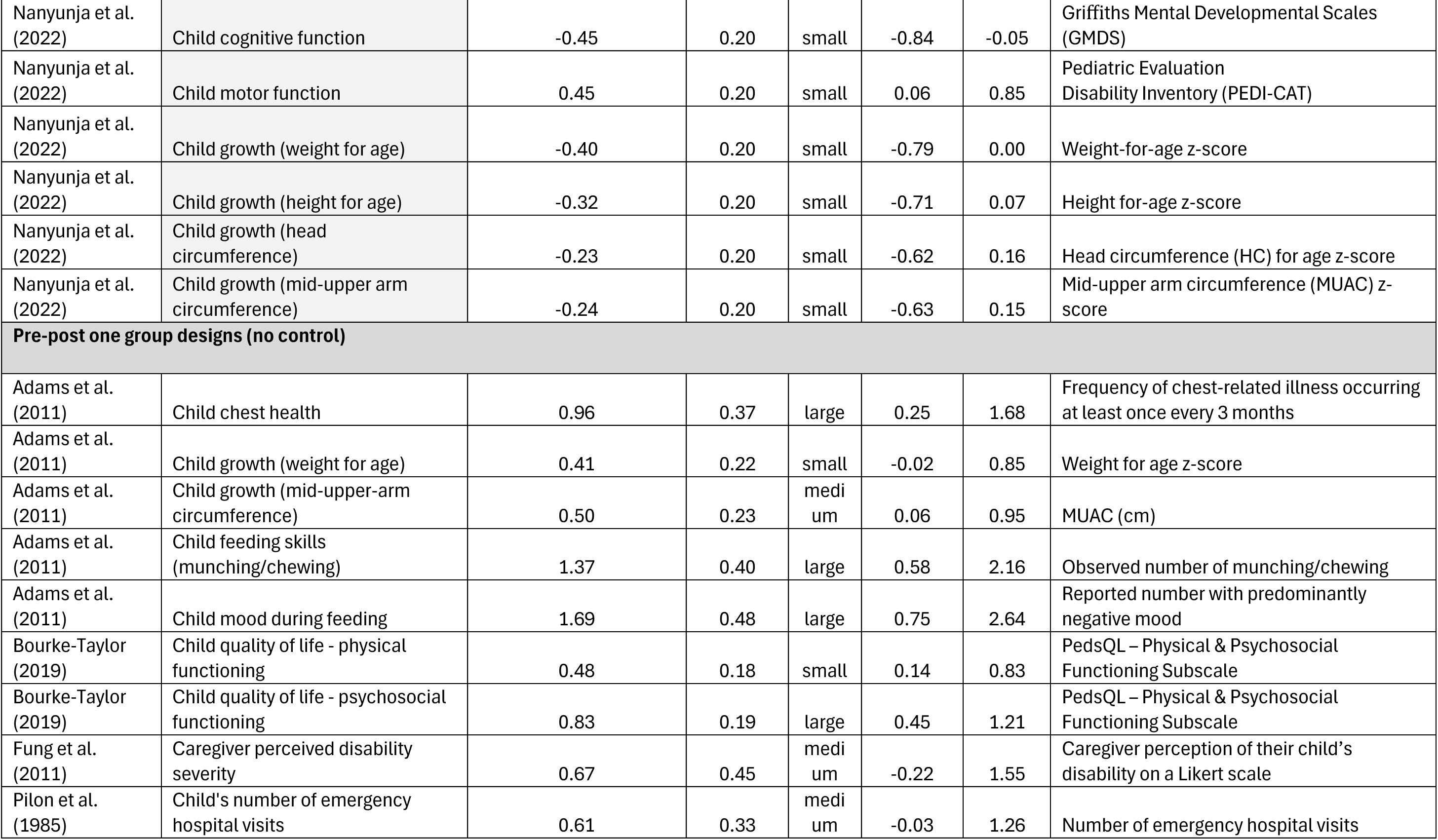
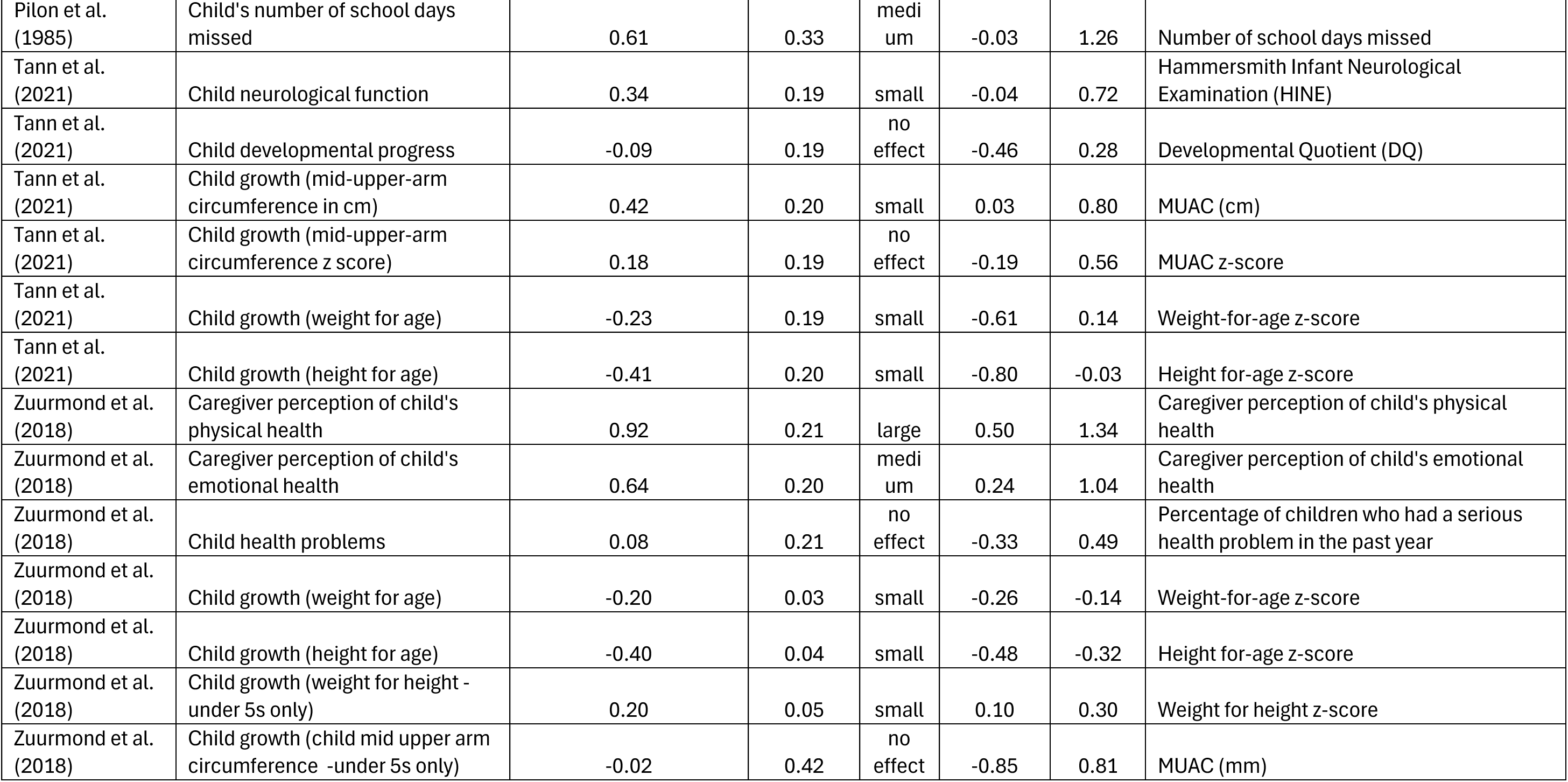
Child-related outcomes.

| Study | Outcome | Standardised Mean Difference (Cohen's d) | Standard Error | Effect size | Lower CI | Upper CI | Measure |
| --- | --- | --- | --- | --- | --- | --- | --- |
| **Independent group designs (intervention and control groups)** |  |  |  |  |  |  |  |
| Al Imam et al. (2022) Arm A | Child quality of life | 0.09 | 0.15 | no effect | -0.21 | 0.39 | TNO-AZL Preschool children Quality of Life questionnaire (TAPQOL) |
| Al Imam et al. (2022) Arm B | Child quality of life | -0.24 | 0.15 | small | -0.54 | 0.06 | TNO-AZL Preschool children Quality of Life questionnaire (TAPQOL) |
| Al Imam et al. (2022) Arm A | Child motor function | 0.04 | 0.15 | no effect | -0.26 | 0.34 | Gross Motor Function Measure (GMFM)-66 |
| Al Imam et al. (2022) Arm B | Child motor function | 0.07 | 0.15 | no effect | -0.23 | 0.37 | Gross Motor Function Measure (GMFM)-66 |
| Asige et al. (2025) | Child motor function | 0.13 | 0.21 | no effect | -0.30 | 0.51 | Gross Motor Function Measure (GMFM)-66 |
| Asige et al. (2025) | Child mobility | 0.11 | 0.21 | no effect | -0.23 | 0.59 | Ugandan version of the Pediatric Evaluation of Disability Inventory (PEDI-UG) |
| Asige et al. (2025) | Child self-care | 0.18 | 0.21 | no effect | -0.23 | 0.59 | Ugandan version of the Pediatric Evaluation of Disability Inventory (PEDI-UG) |
| Asige et al. (2025) | Child social function | 0.20 | 0.21 | small | -0.20 | 0.61 | Ugandan version of the Pediatric Evaluation of Disability Inventory (PEDI-UG) |
| Karim et al. (2021) | Child motor function | 0.35 | 0.17 | small | 0.01 | 0.69 | Gross Motor Function Measure (GMFM)-66 |
| McConachie et al. (2000) | Child skills | 0.26 | 0.39 | small | -0.51 | 1.03 | Independent Behaviour Assessment Scale |
| Mlinda et al. (2018) | Child's mood during feeding | 0.63 | 0.20 | medium | 0.24 | 1.02 | Percentage increase of those rated 'good' vs 'poor' |
| Mlinda et al. (2018) | Child feeding skills (functional skills) | 0.45 | 0.20 | small | 0.07 | 0.84 | Percentage increase of 'yes' vs 'no' |
| Mlinda et al. (2018) | Child feeding skills (oral motor) | 0.16 | 0.19 | no effect | -0.22 | 0.53 | Percentage increase of 'yes' vs 'no' |
| Nanyunja et al. (2022) | Child cognitive function | -0.45 | 0.20 | small | -0.84 | -0.05 | Griffiths Mental Developmental Scales (GMDS) |
| Nanyunja et al. (2022) | Child motor function | 0.45 | 0.20 | small | 0.06 | 0.85 | Pediatric Evaluation Disability Inventory (PEDI-CAT) |
| Nanyunja et al. (2022) | Child growth (weight for age) | -0.40 | 0.20 | small | -0.79 | 0.00 | Weight-for-age z-score |
| Nanyunja et al. (2022) | Child growth (height for age) | -0.32 | 0.20 | small | -0.71 | 0.07 | Height for-age z-score |
| Nanyunja et al. (2022) | Child growth (head circumference) | -0.23 | 0.20 | small | -0.62 | 0.16 | Head circumference (HC) for age z-score |
| Nanyunja et al. (2022) | Child growth (mid-upper arm circumference) | -0.24 | 0.20 | small | -0.63 | 0.15 | Mid-upper arm circumference (MUAC) z-score |
| **Pre-post one group designs (no control)** |  |  |  |  |  |  |  |
| Adams et al. (2011) | Child chest health | 0.96 | 0.37 | large | 0.25 | 1.68 | Frequency of chest-related illness occurring at least once every 3 months |
| Adams et al. (2011) | Child growth (weight for age) | 0.41 | 0.22 | small | -0.02 | 0.85 | Weight for age z-score |
| Adams et al. (2011) | Child growth (mid-upper-arm circumference) | 0.50 | 0.23 | medium | 0.06 | 0.95 | MUAC (cm) |
| Adams et al. (2011) | Child feeding skills (munching/chewing) | 1.37 | 0.40 | large | 0.58 | 2.16 | Observed number of munching/chewing |
| Adams et al. (2011) | Child mood during feeding | 1.69 | 0.48 | large | 0.75 | 2.64 | Reported number with predominantly negative mood |
| Bourke-Taylor (2019) | Child quality of life - physical functioning | 0.48 | 0.18 | small | 0.14 | 0.83 | PedsQL – Physical & Psychosocial Functioning Subscale |
| Bourke-Taylor (2019) | Child quality of life - psychosocial functioning | 0.83 | 0.19 | large | 0.45 | 1.21 | PedsQL – Physical & Psychosocial Functioning Subscale |
| Fung et al. (2011) | Caregiver perceived disability severity | 0.67 | 0.45 | medium | -0.22 | 1.55 | Caregiver perception of their child's disability on a Likert scale |
| Pilon et al. (1985) | Child's number of emergency hospital visits | 0.61 | 0.33 | medium | -0.03 | 1.26 | Number of emergency hospital visits |
| Pilon et al. (1985) | Child's number of school days missed | 0.61 | 0.33 | medium | -0.03 | 1.26 | Number of school days missed |
| Tann et al. (2021) | Child neurological function | 0.34 | 0.19 | small | -0.04 | 0.72 | Hammersmith Infant Neurological Examination (HINE) |
| Tann et al. (2021) | Child developmental progress | -0.09 | 0.19 | no effect | -0.46 | 0.28 | Developmental Quotient (DQ) |
| Tann et al. (2021) | Child growth (mid-upper-arm circumference in cm) | 0.42 | 0.20 | small | 0.03 | 0.80 | MUAC (cm) |
| Tann et al. (2021) | Child growth (mid-upper-arm circumference z score) | 0.18 | 0.19 | no effect | -0.19 | 0.56 | MUAC z-score |
| Tann et al. (2021) | Child growth (weight for age) | -0.23 | 0.19 | small | -0.61 | 0.14 | Weight-for-age z-score |
| Tann et al. (2021) | Child growth (height for age) | -0.41 | 0.20 | small | -0.80 | -0.03 | Height for-age z-score |
| Zuurmond et al. (2018) | Caregiver perception of child's physical health | 0.92 | 0.21 | large | 0.50 | 1.34 | Caregiver perception of child's physical health |
| Zuurmond et al. (2018) | Caregiver perception of child's emotional health | 0.64 | 0.20 | medium | 0.24 | 1.04 | Caregiver perception of child's emotional health |
| Zuurmond et al. (2018) | Child health problems | 0.08 | 0.21 | no effect | -0.33 | 0.49 | Percentage of children who had a serious health problem in the past year |
| Zuurmond et al. (2018) | Child growth (weight for age) | -0.20 | 0.03 | small | -0.26 | -0.14 | Weight-for-age z-score |
| Zuurmond et al. (2018) | Child growth (height for age) | -0.40 | 0.04 | small | -0.48 | -0.32 | Height for-age z-score |
| Zuurmond et al. (2018) | Child growth (weight for height - under 5s only) | 0.20 | 0.05 | small | 0.10 | 0.30 | Weight for height z-score |
| Zuurmond et al. (2018) | Child growth (child mid upper arm circumference -under 5s only) | -0.02 | 0.42 | no effect | -0.85 | 0.81 | MUAC (mm) |

### Certainty of the evidence

The most frequently reported outcomes were assessed for certainty using the GRADE approach (table 6) (60). All outcomes were either ‘low’ or ‘very low’ in the confidence of the effect estimates. Caregiver anxiety/stress and caregiver depression both scored ‘low’ for overall certainty while the rest of the outcomes scored ‘very low’ (caregiver quality of life, caregiver knowledge/skills, family quality of life and family functioning). The high risk of bias in most included studies resulted in downgrading all the outcomes. Caregiver anxiety/stress, depression and quality of life were all consistent in the direction of effects and showed similar effect sizes. These three outcomes were directly related to the research question; however family quality of life and family functioning were seen as more of a proxy outcome and both showed inconsistency in the results. All outcomes were downgraded based on imprecision due to frequently relying on small study numbers and wide confidence intervals.

**Table 6:** GRADE evidence profile for most frequently reported outcomes.

| Outcome | Study design & number | Starting certainty & rationale | Risk of bias | Inconsistency | Indirectness | Imprecision | Other considerations | Overall certainty |
| --- | --- | --- | --- | --- | --- | --- | --- | --- |
| Caregiver anxiety and stress | 7 RCTs, 1 quasi-experimental, 5 pre-post studies | High – mostly RCTs | Downgraded 1 for serious RoB – most studies high risk | Not serious – consistent effects; 2 outliers (but with higher RoB) | Not serious – directly relevant | Downgraded 1 for serious imprecision – small N, wide CIs | No upgrade – no large effect identified; no downgrade – no clear evidence of publication bias. | Low (⊕⊕○○) |
| Caregiver depression | 3 RCTs, 2 pre-post studies | High – mostly RCTs | Downgraded 1 for serious RoB – most studies high risk | Not serious – consistent effects; 1 outlier (but with higher RoB) | Not serious – directly relevant | Downgraded 1 for serious imprecision – small N, wide CIs | No upgrade – no large effect identified; no downgrade – no clear evidence of publication bias. | Low (⊕⊕○○) |
| Caregiver quality of life | 3 RCTs, 3 pre-post studies | Moderate – equal RCT and pre-post studies | Downgraded 1 for serious RoB – most studies high risk | Not serious – most showed small improvement, similar sizes | Not serious – directly relevant | Downgraded 1 for serious imprecision – some no effect or wide CIs | No upgrade – no large effect identified; no downgrade – no clear evidence of publication bias. | Very Low (⊕○○○) |
| Caregiver knowledge and skills | 3 RCTs, 3 pre-post studies | Moderate – equal RCT and pre-post studies | Downgraded 1 for serious RoB – most studies high risk | Not serious – consistent improvements; 1 outlier explained by measurement tool | Not serious – directly relevant | Downgraded 1 for serious imprecision – mixed N and CI widths | No upgrade – no large effect identified; no downgrade – no clear evidence of publication bias. | Very Low (⊕○○○) |
| Family quality of life | 2 RCTs, 4 pre-post studies | Low – mostly pre-post studies | Downgraded 1 for serious RoB – most studies high risk | Downgraded 1 for serious inconsistency – varying sizes; 1 negative effect | Downgraded 1 for serious indirectness – proxy outcome for caregiver wellbeing | Downgraded 1 for serious imprecision – small N, wide CIs | No upgrade – no large effect identified; no downgrade – no clear evidence of publication bias. | Very Low (⊕○○○) |
| Family functioning | 1 RCT, 4 pre-post studies | Low – mostly pre-post studies | Downgraded 1 for serious RoB – most studies high risk | Downgraded 1 for serious inconsistency – varying effects (positive, negative or no effect) | Downgraded 1 for serious indirectness – proxy outcome for caregiver wellbeing | Downgraded 1 for serious imprecision – small N, wide CIs | No upgrade – no large effect identified; no downgrade – no clear evidence of publication bias. | Very Low (⊕○○○) |

*RCT = Randomised controlled trial, RoB= Risk of bias, N= number of participants in study, CIs= Confidence Intervals*

## Discussion

### Key Findings

This systematic review synthesised data on the effects of group programmes targeting improved skills, knowledge and confidence amongst caregivers of children with complex neurodisability, particularly those with a physical impairment from a neurological cause. Twenty-one studies met the inclusion criteria (n=1491) (RCTs =9, quasi-experimental =2, single-group pre-post =10) with 95 outcomes extracted for synthesis and grouped into three caregiver categories; (1) caregiver wellbeing outcomes, (2) caregiver skills and confidence outcomes, and (3) social support and family outcomes, with child-related outcomes (n=43) presented separately. The majority of studies took place in LMICs, and most programmes targeted female caregivers of children with CP under 5 years. The review found varying effects of the interventions, with almost all outcomes trending towards a positive impact, however most were at a high risk of bias due to being self-reported measures with blinding not possible. Intervention characteristics were well described in relation to the goal of the programme, intervention provider, location, dosage and activities included. There was limited reporting of intervention costs and materials required. All interventions involved caregivers either learning or practising skills to care for their child, or skills to look after themselves.

There were three studies that were high quality and the least at risk of bias, all demonstrated significant improvements in outcomes relating to caregiver social capital, wellbeing, quality of life, skills, knowledge and empowerment. The first was an RCT (72) with 251 caregivers of children with CP in Bangladesh who participated in either a microfinance/livelihood and community-based rehabilitation (CBR) programme or CBR alone (both arms were compared with usual care). The programmes in both arms involved caregiver education workshops. Social capital outcome measures (caregiver participation in their community and social networks) improved significantly (d= 1.95, 1.83). The authors confirmed that further caregiver outcomes will be published in the future, and these would be meaningful for this review. The second noteworthy study was an RCT with 94 caregivers of children/young people with CP in Uganda who participated in a multi-component intervention, of which a key part was the caregiver training programme. Medium to large effects were seen in the blinded assessor outcomes of caregiver knowledge (d=1.11), dressing and feeding skills (d= 0.60, 0.87). Self-reported outcomes also demonstrated significant improvement (caregiver quality of life d= 0.89, caregiver burden and stress d=0.92). The final study was a single group pre-post design with 36 mothers of children with disabilities in Australia who participated in a health and empowerment group-based workshop programme (69). The authors demonstrated appropriate methods for controlling for confounding factors and, despite the small sample size, significant improvements were seen in caregiver anxiety (d=0.58), depression (d=0.52), stress (d=0.73), caregiver general wellbeing (d=0.54), caregiver health-promoting activities (d=0.78), and empowerment (d=0.67). It is interesting to note that the first two RCTs did not find significant effects on child outcomes, but the third pre-post study saw improvements in child quality of life psychosocial and physical functioning (d= 0.48, 0.83). All studies were well-reported in relation to study characteristics, intervention characteristics, and methods, and are examples of research with lower risks of bias.

#### Evidence Limitations

The findings from almost all outcomes from the 21 included studies were rated as subject to a high risk of bias due to a variety of factors. Firstly, in the independent group studies, due to the nature of the interventions, participants could not be blinded to group allocation, and outcomes were often self-reported which meant assessor blinding was often not possible either. There was frequent uncertainty in the selection of the reported results due to studies not having published protocols or trial registrations available. Missing data was another difficulty found in most studies as there was often a high proportion of loss to follow up or non-adherence to programmes. Most single group pre-post study designs did not attempt to control for confounding factors and results therefore need to be interpreted with caution as the evidence may be influenced by uncontrolled confounders.

There is a wider issue in the measurement of caregiver-related outcomes following complex, multi-component interventions. Caregiver wellbeing can be negatively influenced by financial difficulties, and restricted time to follow their own interests or work (78,79). Difficulties that caregivers face often move beyond the diagnosis of the child and can relate to external factors such as seeking out appropriate services, coping with discrimination and stigma, and advocating for their child in the community (20). If all these factors can influence outcomes relating to caregiver wellbeing, it is difficult to know whether improvement seen is due to an intervention, and if so which aspect of the intervention had an impact. In relation to the high proportion of non-adherence, it is well known that caregivers of children with complex neurodisability have a high-caregiving burden (80) and their baseline level of wellbeing is often very low (81). These factors can impact a caregiver’s time and capacity to take part in a programme, even if the programme is aimed at improving said factors. Finally, these difficulties can be compounded when caregivers live in poverty as external factors such as transport to the intervention can impact on their ability to participate (82)

#### Comparison with Existing Literature

This review combined group programmes that aimed to improve the psychological wellbeing of caregivers (e.g. Acceptance and Commitment Therapy groups), the caregivers’ knowledge of caring for their child (e.g. Baby Ubuntu groups or nutrition programmes), or the skills to look after themselves (e.g. Healthy Mothers, Healthy Families workshops). There has been mixed findings from similar reviews in relation to the impact of programmes on the wellbeing or quality of life of caregivers of children (42,44,46). Most of the group programmes in this review were found to have a positive impact on caregiver wellbeing, regardless of whether their primary aim was to improve wellbeing or skills. Interestingly, many of the programmes that targeted improving caregiver skills and knowledge often had greater effects on caregiver wellbeing than the programmes specifically aimed at improving the psychological wellbeing. This reflects what parents of children with CP have rated as important in family-centred care in other studies, which is the provision of knowledge about the child (26,39). It also reflects the importance of peer support, regardless of the intervention aim, which has found to be a protective factor for caregiver psychological wellbeing in another systematic review (35). Most studies in this review reported improved social support outcomes (70,72,73), however outcomes relating to family functioning (the daily activities and family relationships in the context of the child’s health condition) were more mixed (69,75,76,83). This relates back to the many external factors that impact a family’s life when they have a child with a disability (20) and raises the question whether an intervention can have an impact on this.

The Family Empowerment Scale (84) was used in two studies in this review describing different programmes in Australia (69) and South India (71) and in both instances, the improved results were statistically significant and showed medium effect sizes (d=0.67, 0.75). These two studies showed improved caregiver wellbeing too. Most studies in this review that reported improved caregiver skills and confidence outcomes also had improved wellbeing outcomes (65,66,69–71,83), This result is echoed by a previous study that demonstrated a link between self-efficacy, a key part of empowerment and confidence, and the mental health of caregivers of children with CP (85).

A few programmes reported no or negative effects but provided rich qualitative data describing the impact on the caregivers’ lives. For example, the ‘Hambisela’ training programme in South Africa helped caregivers to understand what CP was, to accept their children and to know how to care for them appropriately (76) however no effects were found for caregiver quality of life or stress levels. Nanyunja et al. (76) found similar qualitative results with the ‘Baby Ubuntu’ programme in Uganda relating to improved attitudes towards the child, improved wellbeing and confidence, and improved peer support and information sharing, even though these were not reflected in the quantitative data. The online Acceptance and Commitment Therapy group in Australia reported qualitative data relating to parents feeling more mindful, less alone, being more present with family, and accepting their emotions (86). This was not always reflected in the quantitative results relating to mindfulness and stress. These findings echo previous research showing that quantitative tools often miss the true impact programmes have on caregivers. For example, in a UK RCT of the ‘Healthy Parent Carers’ programme, some participants felt the questionnaires failed to reflect their positive experiences (87). Similarly, a recent qualitative study found a widely used wellbeing scale sometimes overlooked important changes, especially negative ones, and did not always reflect caregivers’ lived realities.

Although most studies were published between 2018 and 2023, the fact that a study published in 1985 met the inclusion criteria and demonstrated effectiveness of their group programme in improving outcomes for caregivers and children, demonstrates that this approach is not novel. Caregivers of children with complex neurodisability have been seeking support in the form of information or connections with others in similar situations for decades. Recent literature confirms that caregivers experiences of healthcare services continue to not adequately meet their needs (39,88–93), which perhaps explains why interventions continue to be developed and tested today as they are not yet a part of mainstream services.

### Review Strengths and Limitations

#### Strengths

The effectiveness of caregiver skills training programmes has been widely researched in the field of autism and social communication difficulties, particularly when interventions and outcomes relate to child behaviour (32,94,95). This review clearly maps out different types of caregiver skills training programmes that target children with motor disorders and are delivered in a group format, providing an opportunity for clinicians and researchers to consider various intervention components relevant for implementation in their settings. Many of these programmes were developed in low-resource settings, which may be because community-based, group programmes are more cost-effective, or because task-shifting (96–98) is a necessity due to lack of resources and access to healthcare. This review highlights the innovations taking place in LMICs and provides an opportunity to learn about low-cost, community-based participatory approaches to supporting caregivers of children with complex neurodisability.

#### Limitations

A few studies were not picked up through the search strategy but were instead found through key contacts in the field. This was due to the journals not being indexed in the databases searched. It cannot be ruled out that other studies may have been missed in the same way, although effort was made to hand search reference lists of similar reviews and the included studies, as well as contacting key authors. Another limitation was that studies were excluded if they were not published in English, for example a study in Korea examined the effects of a group empowerment program on mothers of children with CP (99), which would have been relevant to this review, but the full-text was not available in English.

As most of the studies reported complex interventions with multiple components, it was difficult to tease apart which components of the programmes were critical for improved outcomes. For example, a family-centred village-based early intervention programme in India for 135 children with disabilities (including a caregiver intervention) (71) resulted in significant improvements in caregiver empowerment (d=0.75) and reduced caregiver stress (d=0.70). The programme included therapy for the children as well as parent groups, and it is impossible to know which aspects contributed to the improved empowerment and stress. Future reviews on this topic may benefit from using Qualitative Comparative Analysis to identify key components of interventions that lead to success (100). Another helpful way to overcome this is to report on qualitative data which can provide context to the results. For example, Zuurmond et al’s study (74,83) described how caregivers attributed improved wellbeing to increased understanding of their child’s condition, feeling more hopeful and positive towards their child, and the social support that the group provided.

### Implications and recommendations

From a wider systematic perspective, the findings from this review demonstrated the vast expertise to be found in resource constrained settings in relation to the development, testing and implementation of group-based caregiver training programmes for children with neurodisability, particularly with diagnoses like CP. This finding suggests that practitioners in HIC settings, where caregivers’ needs are often unmet, could draw on innovations from LMICs and reconsider assumptions that learning cannot occur in this direction (103). It is important to challenge these assumptions as Harris and colleagues’ (104) note; ‘They hear “Africa” and they think they there can’t be any good services’. Another key learning relevant to practice, is that despite the different aims and components of the group-based programmes, impact on outcomes were broadly similar. It may be that being in a group with others who have similar experiences, is more important than the aim or content of the programme itself. Clinicians and other key partners working with families who have children with neurodisability may consider providing their support, in whichever form this may take, in a group setting. Given the limitations and uncertainty of the evidence in this review, it is recommended that future research in this topic consider better reporting of protocols and trial registrations. Researchers should anticipate the characteristics of this population and the risk of high attrition in studies and plan accordingly, for example covering participants’ transport costs. If conducting a single-group pre-post intervention study, consideration needs to be taken in adjusting for confounding factors to reduce the risk of bias and increase confidence in the results. Finally, it is recommended that a core outcome set be developed to effectively capture the impact of such programmes on caregivers.

## Conclusion

This review has synthesised the existing research on group-based programmes to support the skills and wellbeing of caregivers of children with neurodisability presenting with motor impairments, such as cerebral palsy. It complements previous systematic reviews which have focused on similar interventions for caregivers of children with autism, social communication, and behavioural difficulties. Nearly all studies included in the reviews, regardless of the aims or components of the group programmes, had a positive impact on caregiver wellbeing, confidence, skills, social support and family quality of life. However, most outcomes had a high risk of bias and low levels of certainty in the effects, which poses difficulties for interpretation and drawing strong conclusions or recommendations. Future studies should pay attention to adequate statistical power, robust randomised controlled trial methodology, outcome measurement, confounding factors, costs and the use of qualitative methods to explore quantitative effects.

## Supporting information

PRISMA checklist

## Data Availability

All data produced in the present work are contained in the manuscript

## Declarations Ethical Approval

Not applicable

## Acknowledgments

The first author (KP) was funded by the HARP PhD Programme to conduct this research.

AH is supported by the National Institute for Health and Care Research ARC North Thames. The views expressed in this publication are those of the authors and not necessarily those of the National Institute for Health and Care Research or the Department of Health and Social Care or Health.

MH is supported by the NIHR Global Health Research Professorial Fellowship (NIHR302422). This research was supported by the National Institute for Health Research (NIHR) Great Ormond Street Hospital Biomedical Research Centre. The views expressed are those of the authors and not necessarily those of the National Health Service (NHS), the NIHR or the UK Department of Health. The funders had no role in study design, data collection, analysis, or interpretation, or preparation of this manuscript.

## Author Contributions

KP conceptualised the study and conducted the literature search. KP, SD, and DN screened articles and performed risk of bias assessments. KP led the analysis and synthesis. AH, MH, KB and CH co-supervised all stages of the project. KP drafted the manuscript with input, from AH, MH, KB, and CH. All authors reviewed and approved the final manuscript.

